# Enhancing global preparedness during an ongoing pandemic from partial and noisy data

**DOI:** 10.1101/2022.08.19.22278981

**Authors:** Pascal Klamser, Valeria d’Andrea, Francesco Di Lauro, Adrian Zachariae, Sebastiano Bontorin, Antonello di Nardo, Matthew Hall, Benjamin F. Maier, Luca Ferretti, Dirk Brockmann, Manlio De Domenico

## Abstract

As the coronavirus disease 2019 (COVID-19) spread globally, emerging variants such as B.1.1.529 quickly became dominant worldwide. Sustained community transmission favors the proliferation of mutated sub-lineages with pandemic potential, due to cross-national mobility flows, which are responsible for consecutive cases surge worldwide. We show that, in the early stages of an emerging variant, integrating data from national genomic surveillance and global human mobility with large-scale epidemic modeling allows to quantify its pandemic potential, providing quantifiable indicators for pro-active policy interventions. We validate our framework on worldwide spreading variants and gain insights about the pandemic potential of BA.5, BA.2.75 and other sub- and lineages. We combine the different sources of information in a simple estimate of the pandemic delay and show that only in combination, the pandemic potentials of the lineages are correctly assessed relative to each other. Country-level epidemic intelligence is not enough to contrast the pandemic of respiratory pathogens such as SARS-CoV-2 and a scalable integrated approach, i.e. pandemic intelligence, is required to enhance global preparedness.

## Introduction

The coronavirus disease (COVID-19) outbreak, caused by the SARS-CoV-2 virus and first detected in China in early 2020, likely originated from the Huanan seafood wholesale market in Wuhan [1] and continues to spread worldwide. It has forced national governments to pursue country-level elimination strategies [2, 3, 4] or mitigation policies relying on both non-pharmaceutical interventions (NPI) – e.g., physical distancing, wearing masks, hand hygiene, limit large gathering of people, curfews and, in the worst cases, lockdowns [5] – and pharmaceutical ones, such as massive vaccination campaigns and antiviral therapies [6, 7, 8]. Early strict interventions have been shown to be more effective than longer moderate ones in containing national outbreaks in curbing epidemic growth [9], for similar intermediate distress and infringement on individual freedom [10].

In contrast to policy during the early stages of the pandemic, when pharmaceutical interventions were not yet available, most current national efforts to control the virus rely on reactive strategies which alternate enhancement and lifting of NPIs, with the ultimate goal of prevention, or reduction, of pressure on national health systems. To achieve successful containment, such reactive strategies require high capacity for testing and sequencing to continuously monitor the potential emergence of novel viral strains of SARS-CoV-2, whose mutations might be responsible for more severe and/or more transmissible variants with pandemic potential [11]. We define pandemic potential as the ability of a variant to escape population immunity acquired by vaccination or previous infections and to quickly spread worldwide.

Although the emergence of within-host variants with immune escape is likely to be relatively rare [12], sustained community transmission might favor it. When a new variant emerges, it is crucial for policy and decision-making to characterize novel mutations [13, 14, 15], estimate the growth advantage of the new variant with respect to the existing ones [16] and quantify the effectiveness of currently available vaccines [17, 18]. Consequently, any delay in identifying an emerging variant and in determining its key epidemiological parameters introduces uncertainties in the timeline of community transmissions and imported cases which limit, if not completely prevent, effective mitigation responses to take place, similarly to the cryptic transmission of the wild type SARS-CoV-2 which led to the first COVID-19 wave [19]. Combined with limited testing capacity, porous travel screening [20] – at national and, overall, cross-national levels, where international travel play a significant role to amplify the pandemic potential [21, 22, 19] – and lifting of national NPI, the same delays might seriously hinder the timely detection of an emerging variant. The COVID-19 pandemic has been characterized by the regular emergence of such variants [23]. Three important questions arise during the early stages of such a variant, at which point data is missing and noisy: i) can we reconstruct its geographical origin? ii) can we estimate how long it has been spreading undetected in that location? iii) can we quantify the risk of importation to other locations?

In this work, we devise a protocol to quantitatively answer these questions. We show that, by integrating phylogenetic, epidemiological and behavioral analyses within a framework for data-driven and model-informed pandemic intelligence, it is possible to quantify the pandemic potential of an emerging variant and predict the dynamics of subsequent national outbreaks with satisfactory precision. Finally, we propose a simple combination of the different sources of information to qualitatively compare the lineages according to their pandemic delay and find that only the combined measure can reproduce the observed differences.

## Results

### Blueprint for a pandemic intelligence framework

Reliably quantifying the pandemic potential of an emerging variant requires data, and acquiring data requires time. Between the time *t*_0_ of the first undetected case and the time *t*_1_ of the first reported case and its subsequent lineage designation at time *t*_2_, an emerging variant can silently spread within its country of origin and beyond. For example, let us consider the B.1.1.529 lineage of the Omicron variant (also known as BA.1). This was first reported by genomic surveillance teams in South Africa and Botswana on November 25th 2021. Priority actions have been established by the World Health Organization (WHO) for member states on November 26th, with designation as a variant of concern (VOC) [24] required to raise the level of international alert (*t*_3_). By December 16th 2021, there were several reports of an estimated reduction in both vaccine effectiveness against infection and severe disease [25, 26, 27, 18, 28], together with characterisations of the epidemiology of the variant in South Africa [29], Denmark [30] and Norway [31]. Early phylogenetic analysis placed *t*_0_ during the third week of October 2021, about one month before *t*_1_. Three weeks later it had been identified in 87 countries [29].

Figure 1 summarizes this timeline for B.1.1.529, while highlighting the main analytical steps required to define a self-consistent protocol to characterize the pandemic potential of an emerging variant (see Supplementary Fig. S1 for more mechanistic scheme). Figure 1B illustrates how genomic surveillance data and epidemic modeling can be used to infer the spatio-temporal coordinates of the variant’s origin, thus providing information on *t*_0_. This information is used to estimate the importation risk for all countries in the world due to cross-national human flows. Finally, imported cases are used as seeds for community transmission leading to country-level outbreaks, while accounting for the epidemiological parameters characterizing the new variant. Unavoidable uncertainties about *t*_0_ and epidemiological parameters are propagated through the workflow. Plausible scenarios are presented, accounting for distinct levels of case under-reporting in each destination country (Fig. 1C).

**Figure 1:**
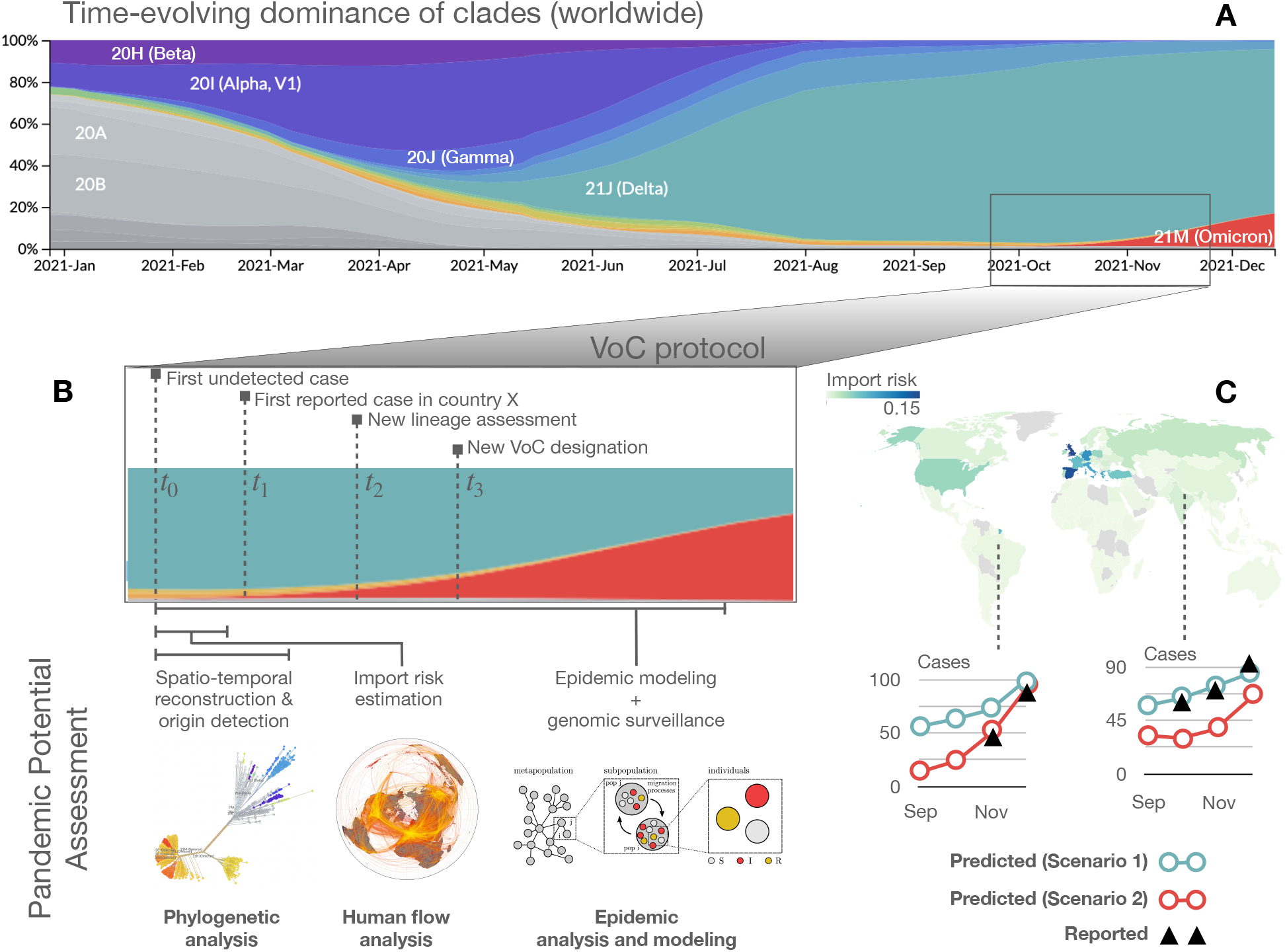
Schematic illustration of our pandemic intelligence workflow. (A) Evolutionary dynamics of SARS-CoV-2 variants, coded by colour. The panel is obtained from `nextstrain.org`, based on GISAID data. (B) For the B.1.1.529 lineage (or BA.1, *Omicron*, according to the WHO nomenclature), we identify four distinct time points in the process of characterising the variant, from the time of the first undetected case to the designation as Variant of Concern. This illustrates how genomic surveillance data is used in combination with global human movement data and epidemic modeling to: i) perform a spatiotemporal reconstruction of the patient zero to identify the country of origin of an emerging variant and estimate its epidemiological parameters and ii) calculate the importation risk for all other countries worldwide. (C) For a subset of about 50 countries worldwide (depending on sequencing data availability), we forecast the increase in the number of cases due to the consequent community transmission according to what-if scenarios, accounting for distinct levels of under-reporting. For a more mechanistic workflow scheme see Supplementary Fig. S1.

In the following, we describe each step of the procedure, detailing our pandemic intelligence framework and the underlying modeling assumptions.

### Reconstructing the origin of an emerging variant in space and time

For all SARS-CoV-2 sequences belonging to the B.1.1.7 (Alpha), B.1.617.2 (Delta), B.1.1.529, BA.2, BA.5, and BA.2.75 (Omicron) lineages from GISAID [32, 33, 34], we retained only those generated from cases reported during the early stage of the corresponding wave from the country of evolutionary origin, from 20 up to a total of 100 sequences per lineage. Where there were multiple candidate countries of origin, we estimated the outbreak country by a simple trait model. We then generated 3 alignments, comprised of respectively 20%, 50% and 100% of the sequence set. These were subsequently cleaned by trimming the 5^′^ and 3^′^ untranslated regions and gap-only sites. Bayesian evolutionary reconstruction of the dated phylogenetic history [35] was used to obtain posterior distributions of the growth rate *t*, the parameters of the molecular clock, and the time of the most recent common ancestor (tMRCA). See Materials and Methods for details.

In this way we obtain an estimate of *t*_0_, the time of the first unreported case, as well as of other epidemic parameters such as the growth rate. From these we estimated the effective reproduction number and generation interval. Indicating the number of infected individuals and number of deaths at time *t* by *I*(*t*) and *D*(*t*) respectively, we consider the time period during which there is co-circulation of an existing variant *v* and an emerging one *ω*. We approximate the epidemic evolution by

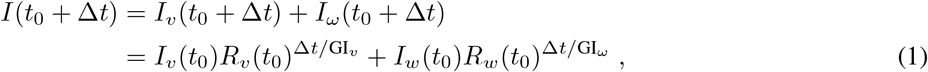

where *I*_*x*_(*t*) is the number of infections due to variant *x* at time t, *R*_*x*_(*t*_0_) is the effective reproduction number at time *t*_0_, and GI_*x*_ is the generation interval. Similarly, the deaths due to the co-circulating variants are approximated by *D*(*t*) = *D*_*v*_(*t*) + *D*_*ω*_(*t*), with

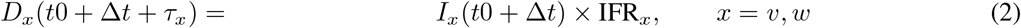

where IFR_*x*_ denotes the infection fatality rate of variant *x* and *τ*_*x*_ is the time lag between infection and death. To fit the unknown epidemiological parameters, i.e. the ones related to variant *ω* for which we obtain a joint probability distribution, we use an optimization procedure (see Materials and Methods).

In the case of B.1.1.529, we obtain *t*_0_ = 29 October 2021 (95% HPD: 20 October–5 November) and a daily growth rate estimate of 0.566 (95% HPD: 0.117–1.035) from the phylogenetic analysis and *t*_0_ = 19 October 2021 (95% CL: 15 October–23 October) from epidemic modelling, with *R*_*t*_ = 2.56 (95% CL: 2.16–3.19) and GI = 7.36 (95% CL: 6.12–9.17). Our results are in good agreement with the literature, reporting *t*_0_ = 9 October 2021 (95% HPD: 30 September–20 October), exponential growth rate of 0.137 (95% HPD: 0.099–0.175) per day [29] and GI = 6.84 days (95% credible intervals: 5.72–8.60) [36].

For further details, refer to Materials and Methods and Supplementary Fig. S2.

### Estimating the importation risk of an emerging variant by country

We use monthly seat capacities of flights between airports from the Official Airline Guide [37], encoding how many people could have travelled if all seats were occupied on flights from airport A to B in the month of the estimated *t*_0_. We indicate the corresponding flow matrix by **F**, where entry *F*_*ij*_ describes the maximal passenger flow to *i* from *j*. The travelling population in the catchment area of an airport is obtained by *N*_*i*_ = *F*_*i*_, with 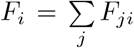, i.e., we assume that the population in the catchment area of the airport is equal to the airports outflow. For each emerging variant, the resulting large-scale network of international travels corresponding to the month of *t*_0_ is used. The import risk is calculated as in [38]: based on the effective distance graph [22] with

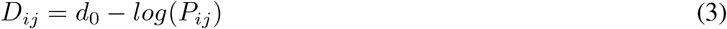

where *P*_*ij*_ is the transition probability from *j* to *i* and a random walk with an exit option, we estimate how likely it is that an infected individual from the emergent variant’s outbreak country reaches any airport worldwide. To work at country level, we aggregate the import risk of all airports of the outbreak country by computing the mean import risk weighted by the international outflux of each airport in the outbreak country. We have performed an extensive analysis to validate the estimated import risk against available data, such as the official arrival times as obtained from the WHO, for each emerging variant. We find considerable correlation between arrival time and import risk distance for different variants (Alpha, Beta, Delta, Gamma) with a median of *r* = 0.55 (range *r* ∈ [0.41, 0.56]). This median is the largest we found, when compared with several alternative distance measures (see Supplementary Figs. S3-S5). The stochastic nature of the reported variant arrival times (based on the by-country rate of genome sequencing, the probabilistic distribution of infected individuals among passengers, etc.) is one possible reason for the imperfect correlation, but another possibility is that the assumed outbreak location is incorrect. To test this, we attempted to identify outbreak locations by recomputing the correlation for all countries (similarly to [22]). For Beta, Gamma, and BA.1 the country declared by the WHO as the outbreak source had the greatest degree of correlation. For Delta and Alpha the WHO candidate had the 2nd and 5th best correlation respectively (see Supplementary Figs. S6, S7). We extended the analysis to sublineages of Omicron and previously circulating variants of interest (VOIs) by estimating arrival times and outbreak countries from GISAID data (see Material and Methods). For 13 of 17 variants the suspected outbreak location from GISAID had at least the 3rd-largest correlation coefficient (of 183), and for all variants the GISAID candidate was at least on the 12th rank (see Supplementary Figs. S8,-S10).

### Modeling country-level epidemic spread of an emerging variant under distinct scenarios

We use results from the previous step of the pipeline as inputs for an epidemic model in order to forecast the potential surge in cases due to an emerging variant in a target country. First, we estimate the daily number of infected people (seeds) traveling to the target country from the country where the VoC emerged (source country), based upon four elements: 1) results of our phylogenetic analysis, which inform both the growth rate and the time of emergence of the variant of concern, 2) genomic surveillance in the source country, 3) estimates of prevalence in the source country (incoporating underreporting), and 4) the import risk score of the target based on estimates from our analysis. Then, we produce short term estimates of the daily incidence of the VoC in the target country by means of a Renewal process [39, 40, 41], in which we take into account both the introductions of seeds from the source country and the local epidemic dynamics caused by secondary cases. The renewal equation approach comes with three main advantages with respect to other models, such as SIR [42]. In fact, 1) it does not require to include in the dynamics the immunological status of the population in the target country; 2) the VoC dynamics can be considered as independent from the ones of the co-circulating VoCs, thus avoiding the need of estimating additional parameters for concurring spreading processes; 3) the model explicitly includes the most relevant epidemiological observables, such as *R*_*t*_, the serial interval distribution [43], and the immune escape of the VoC. For further details we refer to Materials and Methods and Supplementary Figs. S11-S12.

### Assessing the pandemic potential of emerging variants

In Fig. 2 we show the result of each step described above in determining the genomic and epidemiological parameters of the BA.1 lineage and, accordingly, quantify its pandemic potential. We refer the reader to Supplementary Figs. S13-S14 for a more detailed analysis of errors in these estimates. Figure 2A displays a time-resolved maximum clade credibility phylogeny of the lineage. Panel B is the map of import risk across the world. Panels C and D show, for two example countries, the simulated epidemic projections, plotted as weekly incidence. For each reproduction number, the shaded area represents the interval between the estimates derived using the minimum and maximum values of underreporting in the source country. Panel E provides model estimates of case counts in all considered countries.

**Figure 2:**
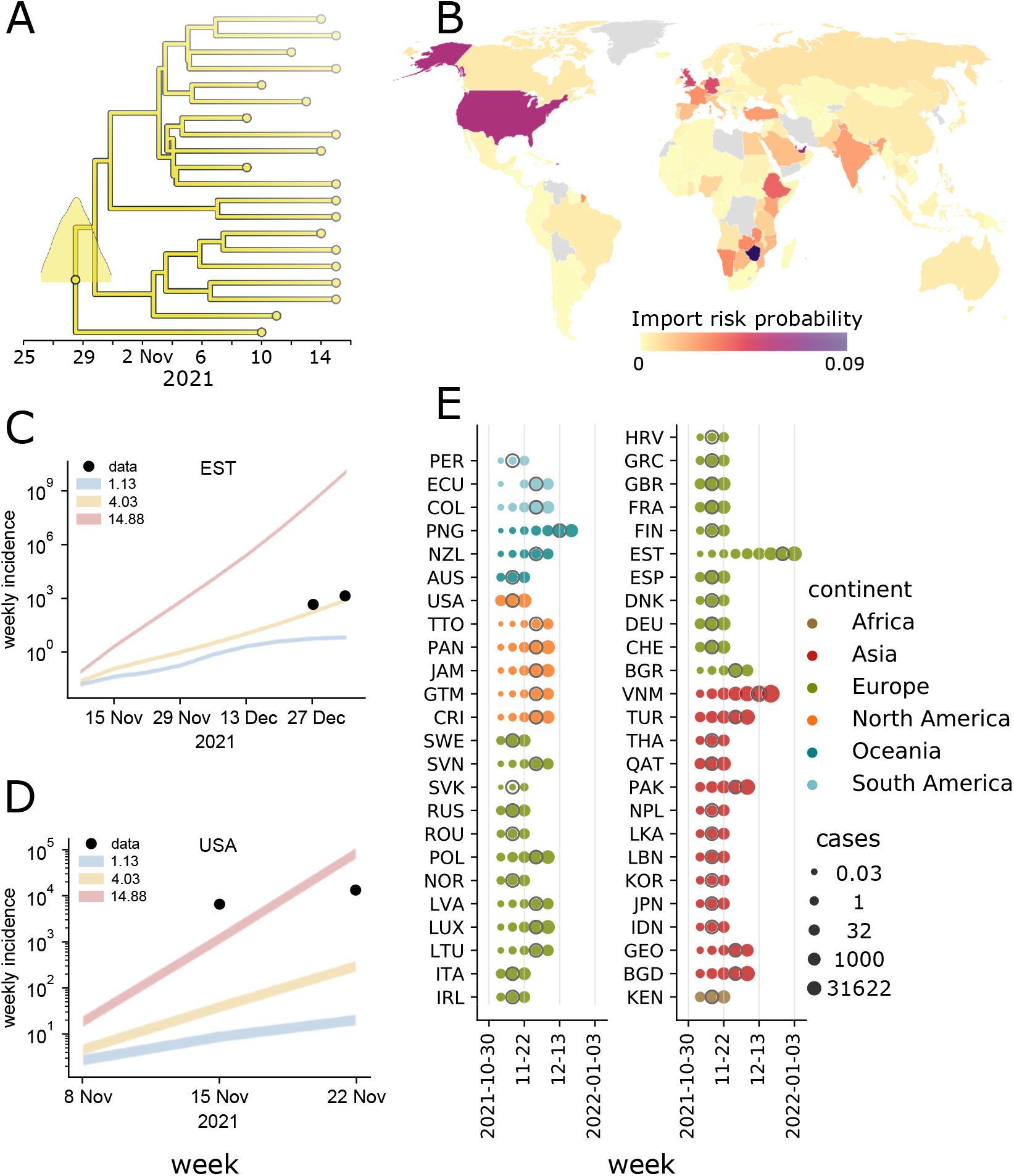
Quantifying the pandemic potential of the B.1.1.529 lineage. (A) Phylogenetic reconstruction and estimation of the most recent common ancestor (MRCA), identified South Africa on 28 October 2021 (95% HPD: 20 October–5 November) as the most likely MRCA. (B) Import risk map: countries are colored by their probability to import infectious individuals carrying the B.1.1.529 (Omicron BA.1) lineage. (C, D) Projected weekly incidence in Estonia and the U.S. obtained from epidemic modeling, under different *R*_*t*_ scenarios indicated by coloured lines. Line thickness represents the range between the minimum and maximum assumed values of underreporting in the source country (here South Africa). Points represent the observed incidence. (E) Case counts simulated using the *R*_*t*_ scenario that corresponds to the mean growth rate from the phylogenetic analysis. For each country, the date of the first reported case is indicated with a grey circle.

Figure 3 shows the results obtained for the SARS-CoV-2 lineages B.1.1.7 (Alpha), B.1.617.2 (Delta), B.1.1.529, BA.2 and BA.5 (Omicron). The date of the most recent common ancestors and the growth rate are shown, together with the temporal evolution of the number of expected cases around 50 countries (varies depending on available sequencing data; Alpha: 59, Delta: 55, BA.2: 51, BA.5: 49 countries). Point estimates of the mean and 95% HPD regions are further provided in Table 1.

**Table 1:**
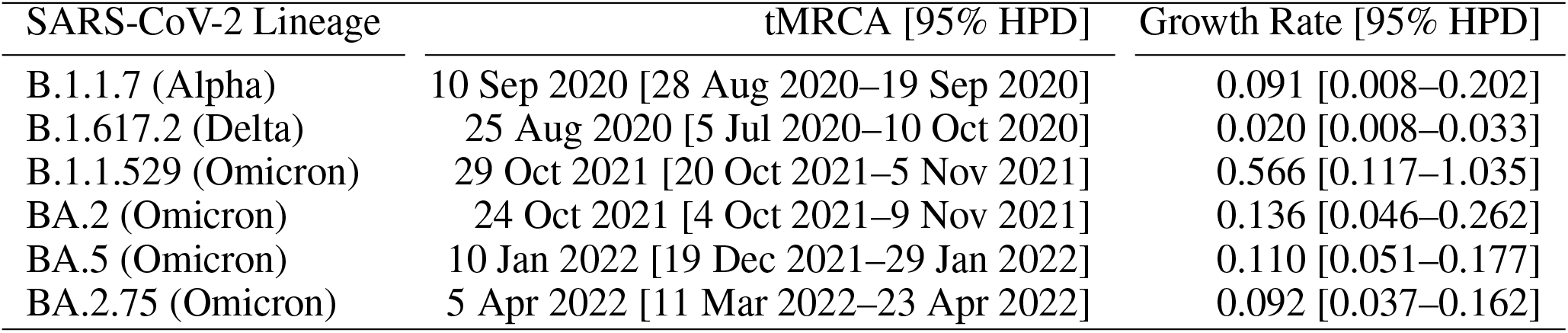
Phylogenetic estimates of the time of most recent common ancestor (tMRCA) and daily growth rate for SARS-CoV-2 B.1.1.7 (Alpha), B.1.617.2 (Delta), B.1.1.529, BA.2, BA.5 and BA.2.75 (Omicron) lineages. Values are expressed as medians and 95% high posterior density intervals.

| SARS-CoV-2 Lineage | tMRCA [95% HPD] | Growth Rate [95% HPD] |
| --- | --- | --- |
| B.1.1.7 (Alpha) | 10 Sep 2020 [28 Aug 2020–19 Sep 2020] | 0.091 [0.008–0.202] |
| B.1.617.2 (Delta) | 25 Aug 2020 [5 Jul 2020–10 Oct 2020] | 0.020 [0.008–0.033] |
| B.1.1.529 (Omicron) | 29 Oct 2021 [20 Oct 2021–5 Nov 2021] | 0.566 [0.117–1.035] |
| BA.2 (Omicron) | 24 Oct 2021 [4 Oct 2021–9 Nov 2021] | 0.136 [0.046–0.262] |
| BA.5 (Omicron) | 10 Jan 2022 [19 Dec 2021–29 Jan 2022] | 0.110 [0.051–0.177] |
| BA.2.75 (Omicron) | 5 Apr 2022 [11 Mar 2022–23 Apr 2022] | 0.092 [0.037–0.162] |

**Figure 3:**
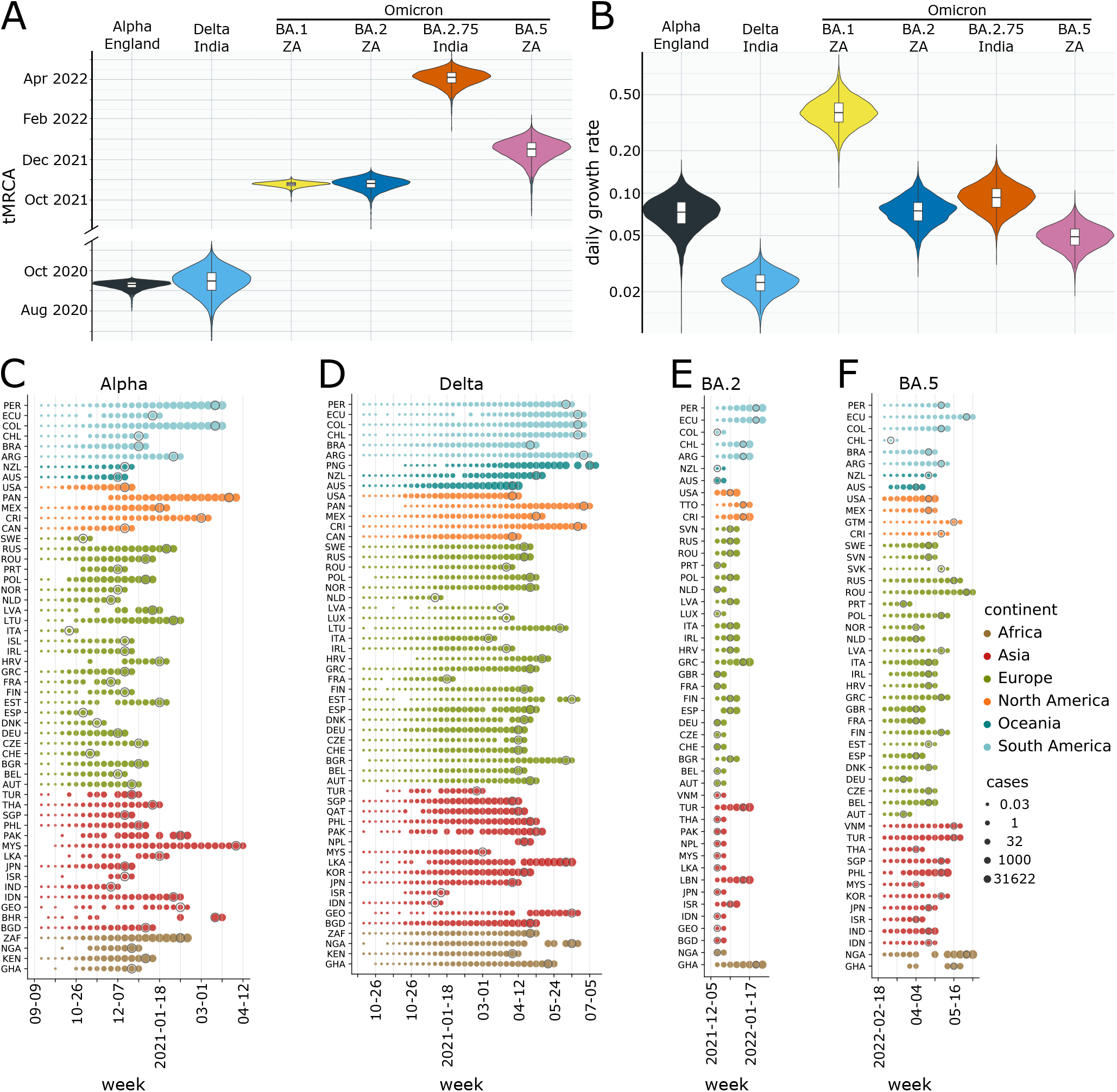
Pan-viral pandemic potential: comparing multiple lineages. (A–B) tMRCA and growth rate estimates for Alpha, Delta, BA.1 (B.1.1.529), BA.2, BA.2.75 and BA.5 from phylogenetic analysis. (C–F) Estimates of case numbers in all the considered countries for the same variants. For each lineage and country, the epidemic simulation starts at the time of infection *t*_0_ of the first undetected case as identified using the phylogenetic analysis. The simulation stops at the third date at which sequences belonging to the considered lineages are greater than zero. Results are provided in logarithmic scale and dates at which the first case is reported are marked with grey circles.

To assess the prediction error of our workflow we computed the normalized root-mean-square error (nRMSE) between prediction scenarios and observations. The nRMSE is zero, if the observation lies in between the simulation scenarios. Otherwise, the nRMSE is the RMSE between observation and the closest prediction scenario, normalized to the range that is spanned by the observations in the respective target country (for details see Material and Methods). Figure 4 captures the absolute and relative frequency of countries according to their nRMSE. Our predictions are in very good agreement (nRMSE = 0) for Alpha in 81.4%, B.1.1.529 in 53.1%, BA.2 in 52.9%, BA.5 in 49% and for Delta in 12.7% of all considered countries. Note that even though Delta has the smallest amount of countries with incidences falling within scenarios prediction, more than 75% of the countries have a nRMSE ≤ 2.5.

**Figure 4:**
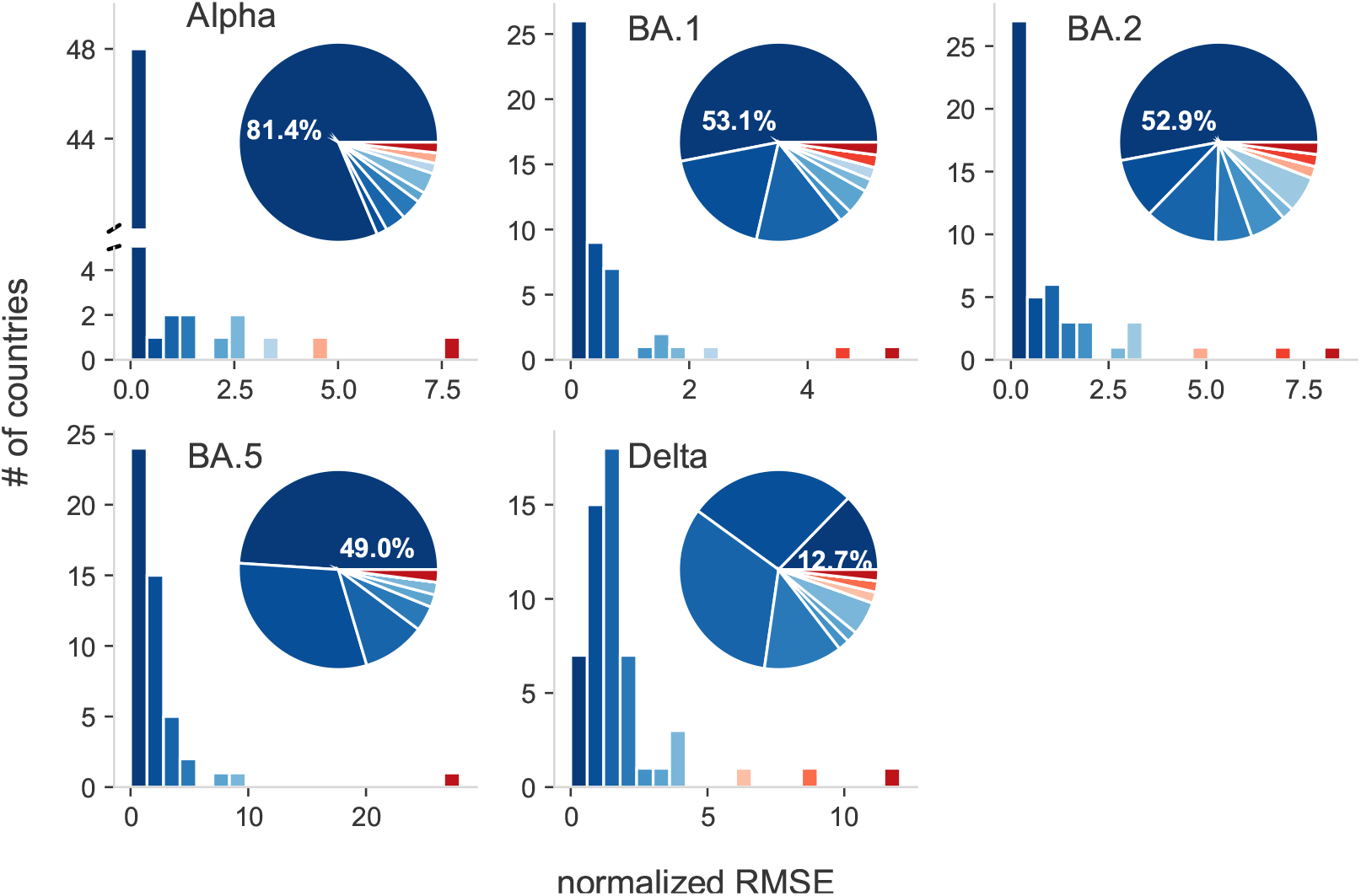
Pandemic intelligence workflow error estimation. Absolute (bars) and relative frequency (segments) of countries according to their normalized root-mean-square error (nRMSE) for the Alpha (B.1.1.7), Delta (B.1.617.2), BA.1 (B.1.1.529), BA.2, BA.5 (Omicron) lineages. The normalized RMSE is zero if the number of infected people evaluated from data is inside the range spanned by the epidemic scenarios. Otherwise, it is the RMSE between observed incidence and the incidence of the closest epidemic scenario, normalized to the range spanned by observed incidences in the respective country. The order and color of the bars and segments is identical, i.e. the bars serves as color legend for the segments. For example, dark blue always represents the number or percentage of countries with the smallest nRMSE.

### The pandemic delay as a simple integrative measure for variant classification

Despite the simplicity of the projection approach, the numerical simulation on country level makes it difficult to summarize an emergent variant’s pandemic potential in simple terms. To close this gap, we introduce the pandemic delay *t*_*y*_, that combines phylogenetic, connectivity and epidemic information in a single equation by assuming that the new variant has a fitness advantage Δ*f* against the preexisting strains and is competing for the infected population estimated via a simple logistic growth equation (see Material and Methods for a detailed derivation). The pandemic delay *t*_*y*_ estimates the time between tMRCA and when the new variant reached a fraction *y* of all sequenced probes in the target country *m*:

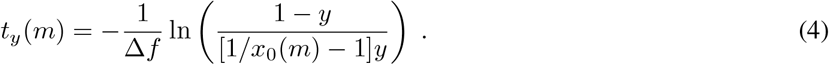

The phylogenetic information is encoded in the fitness advantage Δ*f* = ln *R* − ln 1 with *R* as the phylogenetic estimate of the reproduction number, i.e. we assume that the population behaviorally and/or medically adapted to the preexisting strains resulting in a 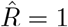. The initial fraction *x*_0_(*m*) encodes the connectivity between outbreak and target country and their epidemic state, i.e. it estimates how many cases are at tMRCA imported relative to the current case number. Figure 5A shows a qualitative agreement between our estimated *t*_*y*_ and the observed pandemic delay 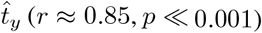 and suggests a linear relation considering all but the Delta lineage’s overestimated delay. Note that also within the lineages, the correlation between estimated and observed delay is in general high and significant ([r-, p-value]: Alpha [0.5, 0.001], Delta [0.3, 0.06], BA.1 [0.52, 0.02], BA.2 [0.14, 0.47], BA.2.75 [0.97, 0.002], BA.5 [0.44, 0.008]), which highlights the importance of the additional connectivity information. The rank correlation between median estimated *t*_*y*_ and the phylogenetic estimate of *R* (Figure 5B) is almost perfect, with the Alpha lineage as an exception that has a shorter pandemic delay (rank 2) than expected if solely *R* would be considered (rank 4) because of the particularly high outflux per capita of its outbreak country (Great Britain). Again, it illustrates that the combination of all information is necessary to gain a realistic estimate of an emergent lineage’s pandemic potential.

**Figure 5:**
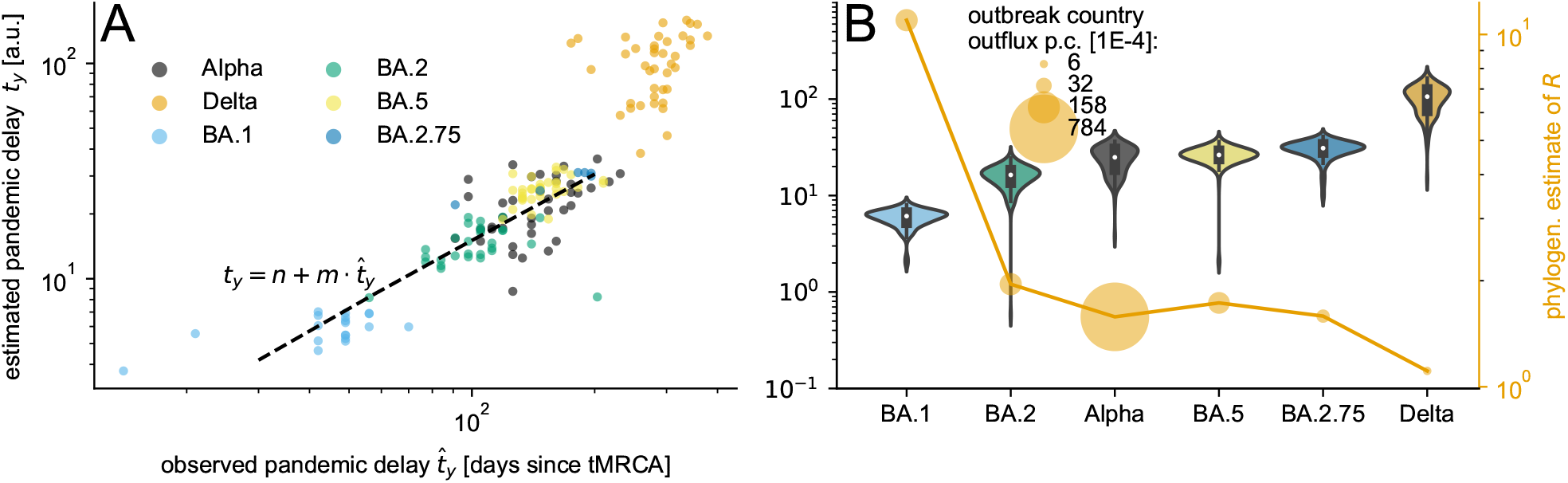
Pandemic delay. The pandemic delay *t*_*y*_ until the lineage reaches a fraction *y* ∈ [0.13, 0.16] is estimated for Alpha, Delta, BA.1 (B.1.1.529), BA.2, BA.2.75 and BA.5. (A) The country specific estimates of *t*_*y*_ versus the observed pandemic delay 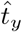 suggest a linear relation, highlighted by a Theil-Sen estimation (dashed line) with slope *m* ≈ 1.55 and intercept *n* = − 0.47 based on all but the Delta lineage data. (B) The lineages sorted according to their median *t*_*y*_ in combination with the phylogenetic estimate of the reproduction number *R* show that the additional connectivity information, the monthly outbreak country outflux per capita (equivalent to the export probability), explains the shorter predicted pandemic delay of the Alpha lineage despite its second-lowest median *R*.

## Discussion and conclusions

We presented an integrated framework that combines phylogenetic analysis of genomic surveillance data with international human mobility data and large-scale epidemic modeling, in order to characterize in nearly real time the pandemic potential of an emerging variant. This concept is intended to provide quantitative indicators about the ability of a variant to escape population immunity acquired by previous infections and/or vaccination, and quickly spread at a global level through human activities.

Our framework naturally deals with missing and noisy information to infer, through a Bayesian approach, the most likely origin – in space (on the country level) and time – of an emerging variant and its growth rate. Spatial and temporal coordinates are used to feed an analytical technique to estimate the probability that a given number of infectious individuals, departing from the country where the variant first appeared, travel to other countries with no exposure to it. This crucial step is based on international travel data, providing information about human movements between countries. Note that our approach is more powerful than naive estimates based only origin-destination pairs: in fact, we make full use of the knowledge we have about the underlying international travel network and its latent geometry [22, 44, 45], known to play a crucial role to amplify the spread of an emerging pathogen [19]. The last stage of our framework is to use importation risk to quantify the number of imported infectious cases to each country and, accordingly, estimate the consequent unfolding of the epidemic due to the emerging variant. The epidemic model is intended to quantify undetected infections that occur well before the first genomic sequence is isolated from a case in a country. Finally we validate our estimate of the pandemic delay, that allows a simple to interpret qualitative comparison between variants incorporating phylogenetic, epidemic and connectivity information. The estimation is based on a logistic growth equation for the relative fraction of a new variant. These predictions will be less accurate if growth advantages in different countries are heterogeneous, for example due to immune escape.

Only the early phase of spread of a new lineage is estimated and the proposed model can safely take advantages of assumptions like a homogeneous mixing and the lack of feedback loops in the epidemic dynamics.

We have validated our integrated framework on existing variants, including B.1.1.7 (Alpha), B.1.617.2 (Delta), B.1.1.529 (BA.1), BA.2, BA.2.75 and BA.5 (Omicron), finding excellent agreement with independent estimates of the relevant phylogenetic and epidemiological parameters. By accounting for different scenarios in the progress of the epidemic in each country, we provide quantifiable indicators to inform decision makers and support pro-active policy interventions to mitigate the potentially harmful effect of an emerging variant. For the current variant of most concern, BA.5, we estimate that its most recent common ancestor existed in early January 2022 (10 January 2022, 95% HPD: 19 December 2021–29 January 2022), with a daily growth rate of 0.110 (95% HPD: 0.051–0.177).

Overall, our findings show that it is possible to aim at pandemic intelligence, even with partial and noisy data. We must caution that the estimates of the pandemic potential of an emerging SARS-CoV-2 variant are largely driven by the uncertainty in the spatio-temporal coordinates of its origin. The Delta lineage is our most unreliable estimate (Figures 3-5), possibly due to the low genomic surveillance at the tMRCA in the outbreak country India, even if Delta and Alpha have a comparable sequencing rate corrected for underreporting (Supplementary Table S1), because the underreporting is based on COVID-mortality that is known to be again underestimated in India by a factor of 6 to 7 [46]. Another reason especially for the overestimation of Delta’s pandemic delay (Figure 5) is its low phylogenetic growth rate estimate, which implies that during the long time till the Delta dominates, additional mutations can happen that potentially speed up the process. Importantly, note that only the validation of our scenario predictions relies on large enough sequencing rates in the target country, but not its application. That means our framework is perfectly suitable for low- and middle-income countries with little to no genomic surveillance, as long as disease related mortality is monitored.

Failures in international cooperation with a view to finding global solutions have undoubtedly shaped the COVID-19 pandemic. We have provided robust evidence that epidemic intelligence at country level is not enough, alone, to contrast the pandemic of respiratory pathogens such as SARS-CoV-2, in the absence of well-coordinated genomic surveillance – especially in low-income and middle-income countries currently lacking and adequate response capacity [47] – and global projections of variant’s pandemic potential. Our approach is inherently integrated and scalable, adding to ongoing modeling efforts and pan-viral analyses [48, 49, 50, 51, 23, 11] and responding to global calls for coordinated action [47, 52, 53]. The data-driven approach provides a vital step in the path towards pandemic intelligence – where the interconnected and interdependent nature of human activities [22, 19, 54] is naturally accounted for at a global level – as well a means of enhancing global preparedness against future emerging variants.

## Data Availability

All data produced in the present study are available upon reasonable request to the authors.

## Acknowledgments

The authors acknowledge the GISAID initiative and all the authors from the originating laboratories where genetic sequence data were generated for sharing such data through GISAID, which has made this work possible.

## Author contributions

M.D.D. designed the study; P.K., V.D., F.D.L., A.Z., S.B., A.D.N., M.H. and L.F. performed the numerical experiments; all authors analyzed the data and wrote the manuscript.

## Data and materials availability

Both the data and analysis material will be available online at Zenodo upon publication. This work is licensed under a Creative Commons Attribution 4.0 International (CC BY 4.0) license, which permits unrestricted use, distribution, and reproduction in any medium, provided the original work is properly cited. To view a copy of this license, visit https://creativecommons.org/licenses/by/4.0/. This license does not apply to figures/photos/artwork or other content included in the article that is credited to a third party; obtain authorization from the rights holder before using such material.

## SI Appendix

**Figure S1:**
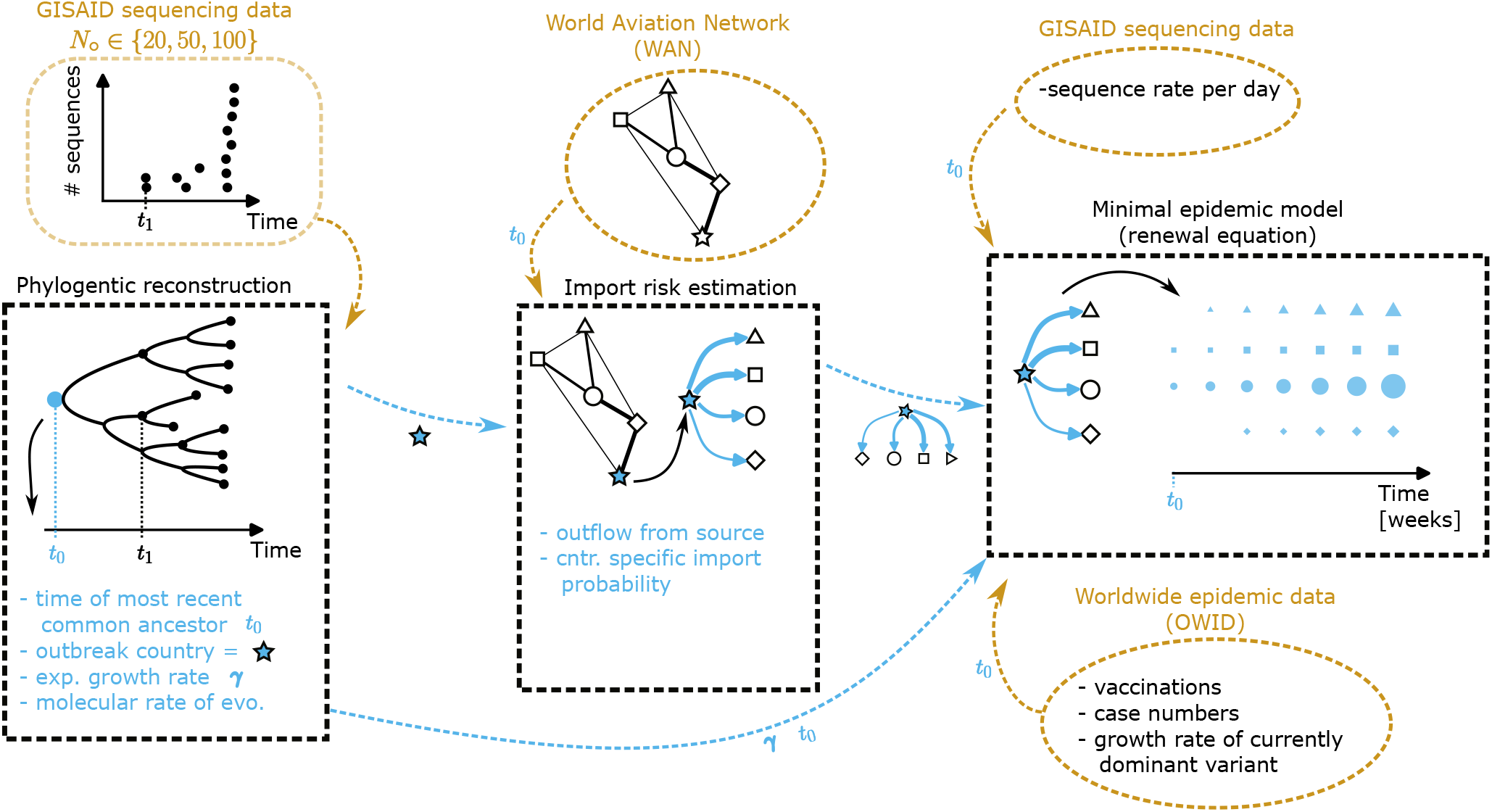
Schematic mechanistic pipeline workflow. The three pieces of our pipeline (black dashed boxes) are illustrated and what input they get from external data sources (orange colored) or from the output of earlier parts of the pipeline (blue arrows). The *t*_0_ close to the orange arrows means that data from the external sources is used at or prior to *t*_0_, which is the estimated time of the most recent ancestor (the output of the first part of the pipeline, the phylogenetic reconstruction).

### Materials and Methods

#### I Phylogenetic Reconstruction

##### I.1 Genomic dataset compilation

We retrieved all SARS-CoV-2 sequences belonging to the Alpha B.1.1.7, Delta B.1.617.2, Omicron B.1.1.529 (BA.1), BA.2, BA.5, and BA.2.75 lineages from GISAID. Each genomic dataset was filtered by only retaining those sequences that were generated from cases reported during the initial wave and from the country of evolutionary origin, up to a total of 100 sequences per lineage. We then generated 3 alignments using MAFFT 7.505 [1], each comprised of 20%, 50% and 100% of the total number of sequences, which were subsequently cleaned by trimming the 5^′^ and 3^′^ untranslated regions and gap-only sites.

##### I.2 Phylogenetic estimates of epidemiological parameters

We performed a common Bayesian evolutionary reconstruction of timed phylogenetic history using BEAST 1.10.5 [2] that was source compiled from its GitHub repository (https://github.com/beast-dev/beast-mcmc). We modelled the nucleotide substitution process according to a *HKY* 85 + Γ parameterisation, setting a strict molecular clock and an exponential growth model as coalescent prior. We used a *Lognormal*(*μ* = 9 × 10^−4^, *σ*^2^ = 1 × 10^−5^) prior for the molecular rate of evolution, a *Laplace*(*μ* = 0, *b* = 100) prior for the rate of exponential growth and a *Lognormal*(*μ* = 5.7, *σ*^2^ = 2.3) prior for the exponentially growing viral population size. We further set an initial calibration for the time of the most recent common ancestor (tMRCA) at an age of ∼ 6 months before the most recent sample included in the alignment. All the remaining priors were left at their default values.

Bayesian inference through Markov chain Monte Carlo (MCMC) was performed for 2 × 10^8^ generations, sampling every 20,000 generations and using the BEAGLE 3.1.2 library to increase computational performance [3]. MCMC convergence and mixing properties were inspected using Tracer 1.7.2 [4] to ensure that effective sample size (ESS) values associated with estimated parameters were all *>*200. After discarding 10% of sampled trees as burn-in, estimates of the growth rate, molecular clock and tMRCA were extracted along with their posterior distributions (Figure S2).

##### I.3 Estimates based on epidemic modeling

We obtain an independent estimate for *t*_0_, the time of the first unreported case, and for other epidemic parameters, such as the effective reproduction number and the generation interval. By indicating with *I*(*t*) the number of infected individuals at time *t* and with *D*(*t*) the number of deaths, we consider the stage with the co-circulation of an existing variant *v* and the emerging one *ω*. Since we consider the final stage of the contagions due to *v* and the early stage of the contagions due to *ω*, we approximate the epidemic evolution by

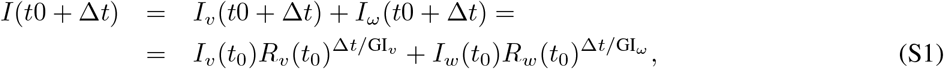

where *I*_*x*_(*t*) is the number of infections due to variant *x* at time t, *R*_*x*_(*t*_0_) is the effective reproduction number at time *t*_0_ and GI_*x*_ is the generation interval. Similarly, the deaths due to the co-circulating variants are approximated by

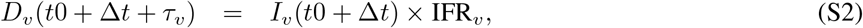

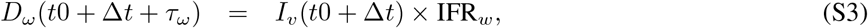

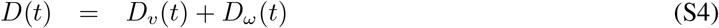

where IFR_*x*_ denotes the infection fatality rate of variant *x* and *τ*_*x*_ is the lag between infection and death. To fit the unknown parameters, i.e. the ones related to variant *ω*, we use particle swarm optimization [5] to minimize the loss function

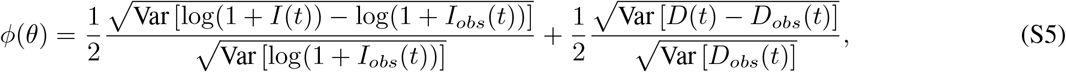

where *I*_*obs*_(*t*) and *D*_*obs*_(*t*) are the number of infected individuals and deaths from empirical data [6], Var indicates the variance in time and *θ* = {*t*_0_; *R*_*ω*_(*t*_0_); GI_*ω*_; IFR_*ω*_; *τ*_*ω*_} is the vector of the epidemiological parameters characterizing the emerging variant, for which we obtain a joint probability distribution.

**Figure S2:**
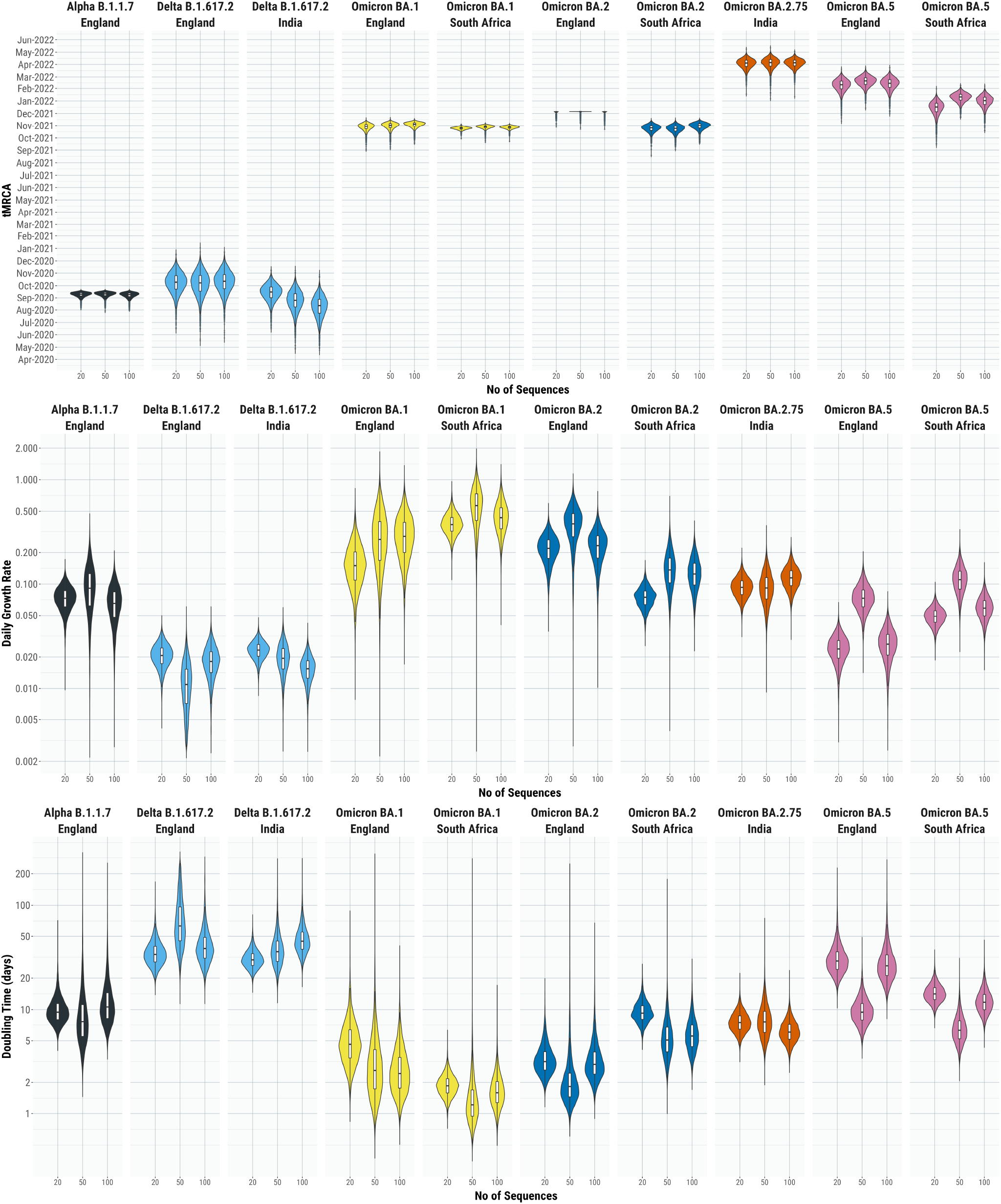
Pan-variant phylogenetic analysis. Posterior distributions of the time of the most recent common ancestor (tMRCA), daily growth rate and doubling time estimated for each of the Alpha B.1.1.7, Delta B.1.617.2, Omicron B.1.1.529 (BA.1), BA.2, BA.5, and BA.2.75 SARS-Cov-2 lineages using alignments of 20, 50 and 100 sequences.

#### II Import Risk estimation

##### II.1 International travel dataset compilation

We retrieve the monthly seat capacities between airports from the OAG (Official Airline Guide). Note, that it does not represent the actual passengers that flew from airport A to B in one month, but the maximal capacity, i.e. how many could have travelled if all seats were occupied. It is therefore an upper limit for the passenger flux and we refer to it as the flow matrix **F**, where *F*_*ij*_ describes the maximal passenger flow to *i* from *j*. We estimate the travelling population in the catchment area of an airport by *N*_*i*_ = *F*_*i*_, with *F*_*i*_ = ∑ _*j*_ *F*_*ji*_, i.e. we assume that the population is proportional to the outflux of the airport. For each variant, we use the WAN at the month of the outbreak day of the respective variant.

**Figure S3:**
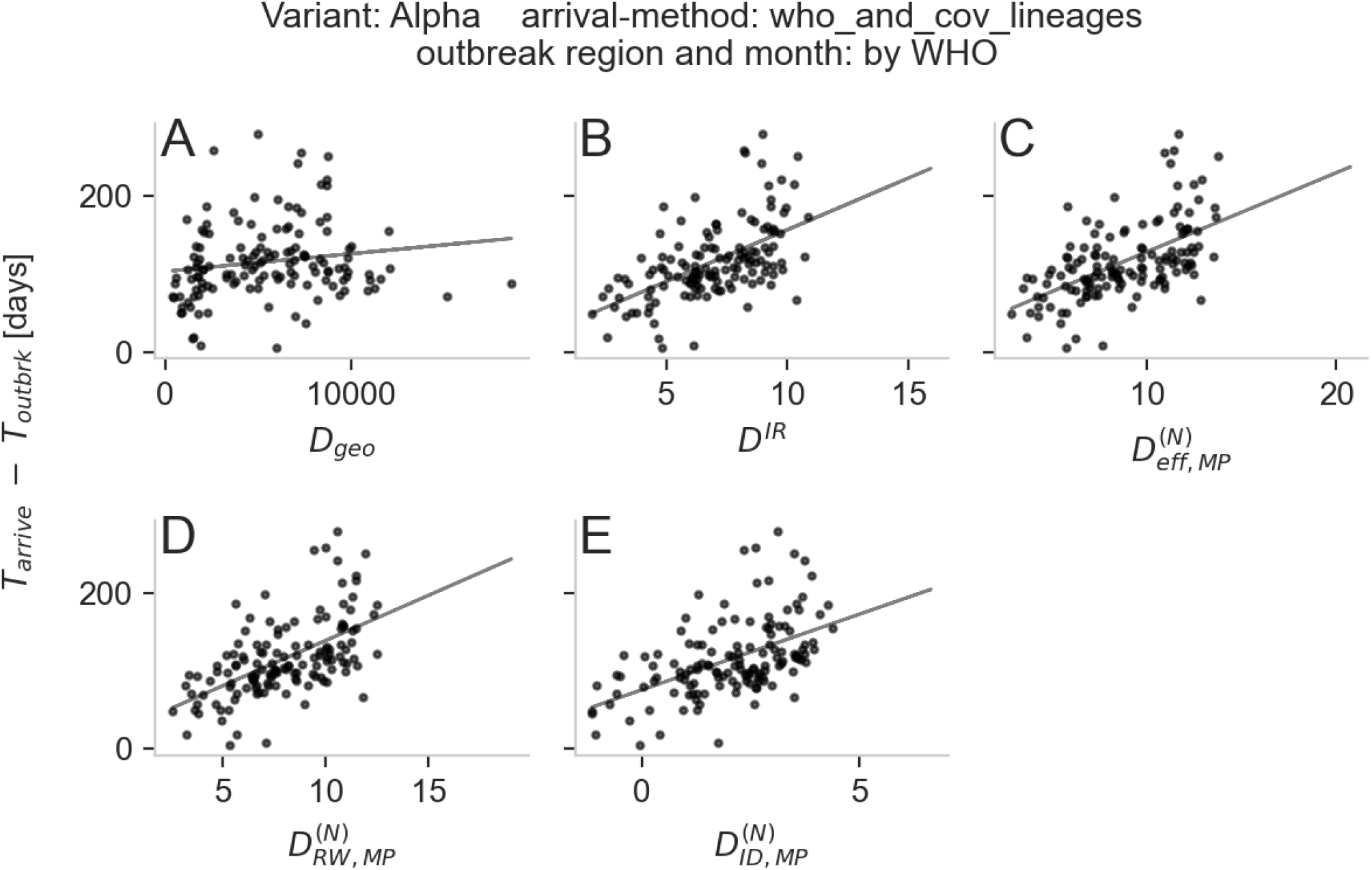
Distance measures vs. arrivals for Alpha variant. The distance measures are the geographic distance *D*_*geo*_ (**A**), the import risk distance *D*^*IR*^ (**B**), the effective distance 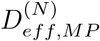 (**C**), the random walk distance 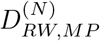 (**D**) and the information diffusion distance 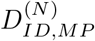 (**E**) whereby the latter three (**C, D, E**) are generalized to weighted multiple paths.

##### II.2 Quantifying the Import Risk

The import risk method is introduced in a separate study [9] where it is compared to another data-driven estimate. Here we present a short outline of the method. To know how many passengers leave at node *j* given they started at node *i*, we introduce the shortest path exit probability *q*(*j*|*i*) (SPEx). It is based on the shortest path tree of the effective distance [10], and combines the exit probability with all possible paths that end in *j*. The resulting import risk is therefore an extension of the SPEx.

In order to compute the **SPEx** we first define, with the flow matrix (maximal passenger flux) **F** and the travelling population of the catchment area *N*_*i*_, the transition matrix **P**, where the element *P*_*ij*_ = *F*_*ij*_*/* ∑_*i*_ *F*_*ij*_ = *F*_*ij*_*/F*_*j*_ is the probability to transition to *i* from *j*. Now, the effective distance graph [10] is *D*_*ij*_ = *d*_0_ − *log*(*P*_*ij*_), with *d*_0_ as the distance offset which we set to *d*_0_ = 1 (the larger *d*_0_ the more *D*_*ij*_ increases with increasing hop-distance). Let **T**(*n*_0_) be the shortest path tree on **D** for the point of origin *n*_0_. With respect to node *n* the downstream nodes Ω(*n*|*n*_0_) are those nodes that can be reached from the source *n*_0_ through node *n* on **T**(*n*_0_).

**Figure S4:**
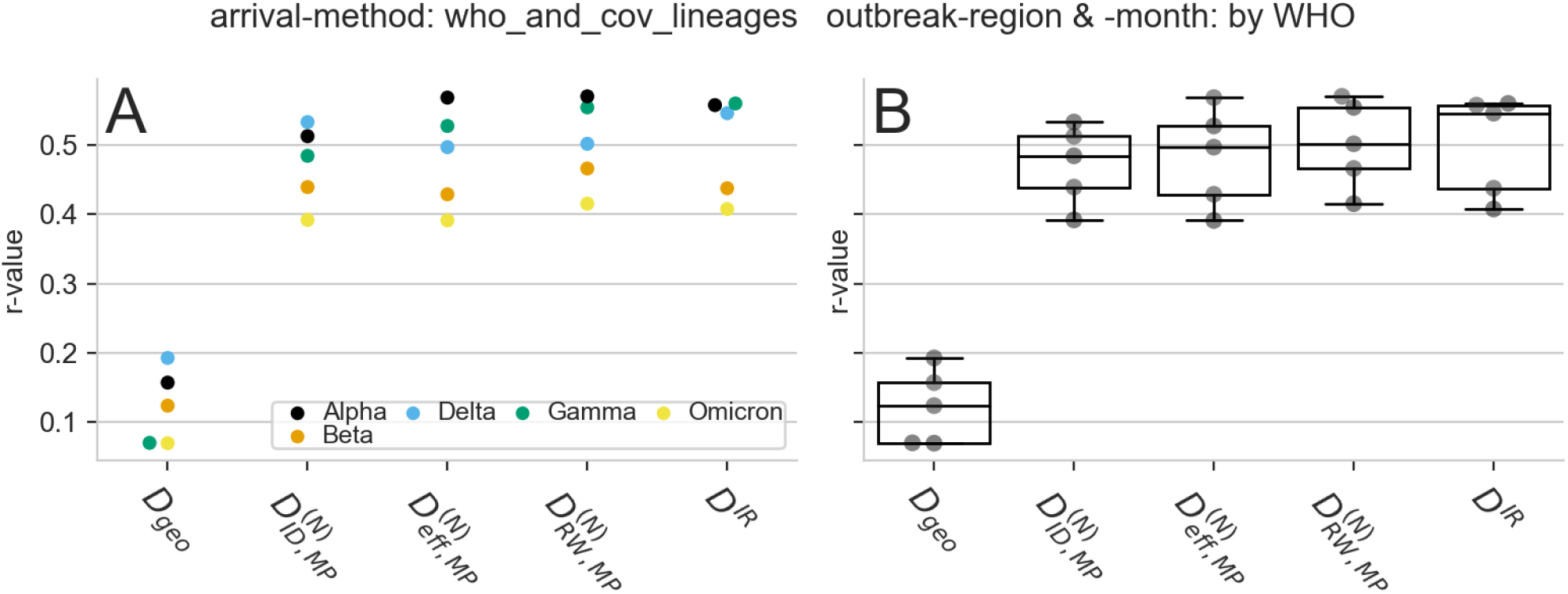
Correlation comparison between different distance measures. The distance measures are the geographic distance *D*_*geo*_, the import risk distance *D*^*IR*^, the effective distance 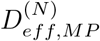, the random walk distance 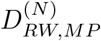 and the information diffusion distance 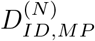 whereby the latter three are generalized to weighted multiple paths.

**Figure S5:**
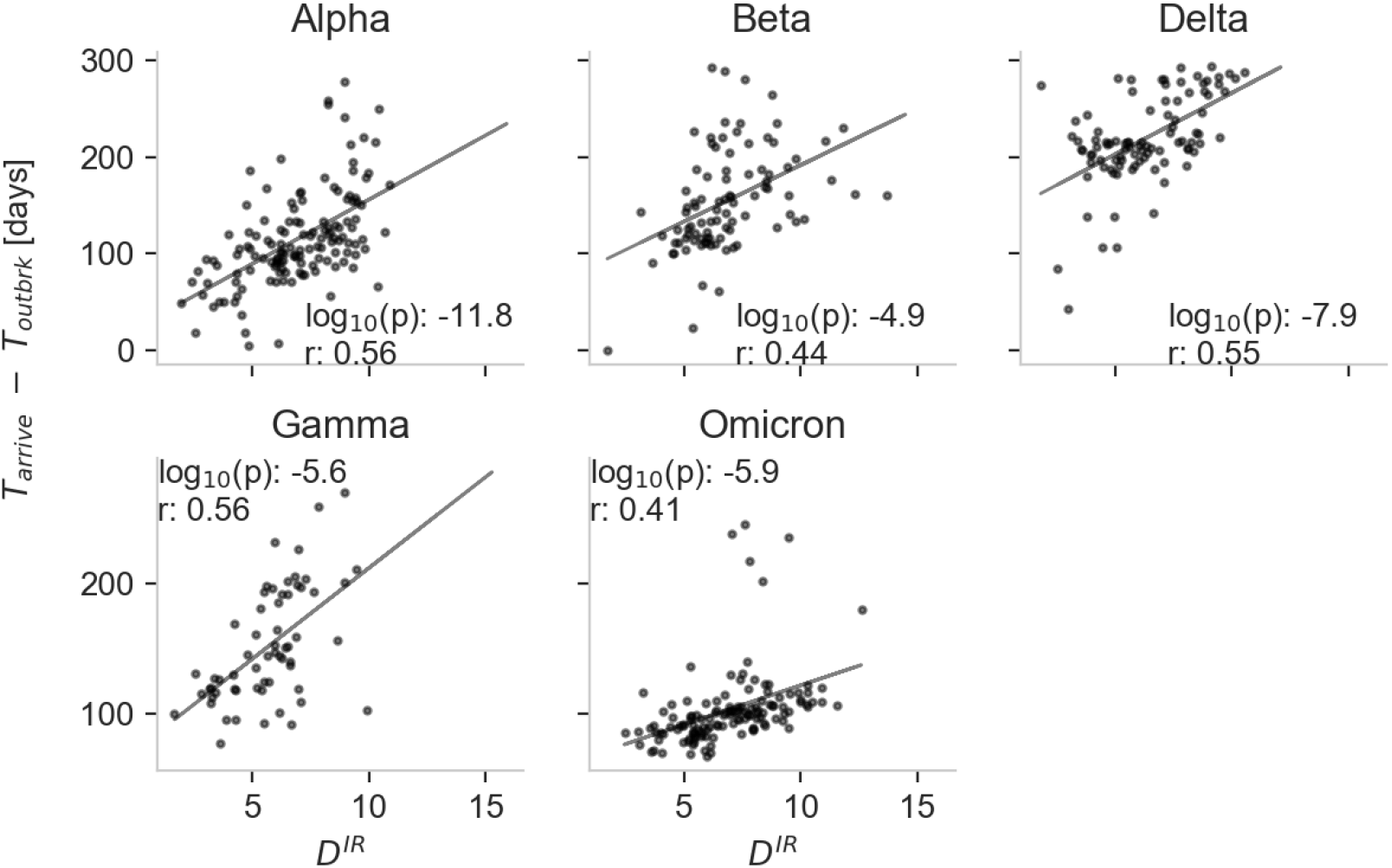
Correlation of arrival times of variants with the import risk distance *D*^*IR*^. For the import risk distance *D*^*IR*^(*m*|*n*_0_) = − log(*p*_∞_(*m* | *n*_0_)) the WAN of the WHO outbreak month is used and the WHO outbreak location as source country. The arrival times are taken from the “cov-lineages.org” [7, 8] project.

Now we compute the SPEx *q*(*i*| *n*_0_) by assuming that all passenger that start at *n*_0_, travel along the shortest path tree **T**(*n*_0_) and distribute to other airports according to their respective populations *N*_*n*_. We assume that the exit probability at *i* is proportional to the ratio of the population at *i* (i.e. *N*_*i*_) to the population of all of *i*’s downstream nodes 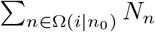 plus *N*_*i*_:

**Figure S6:**
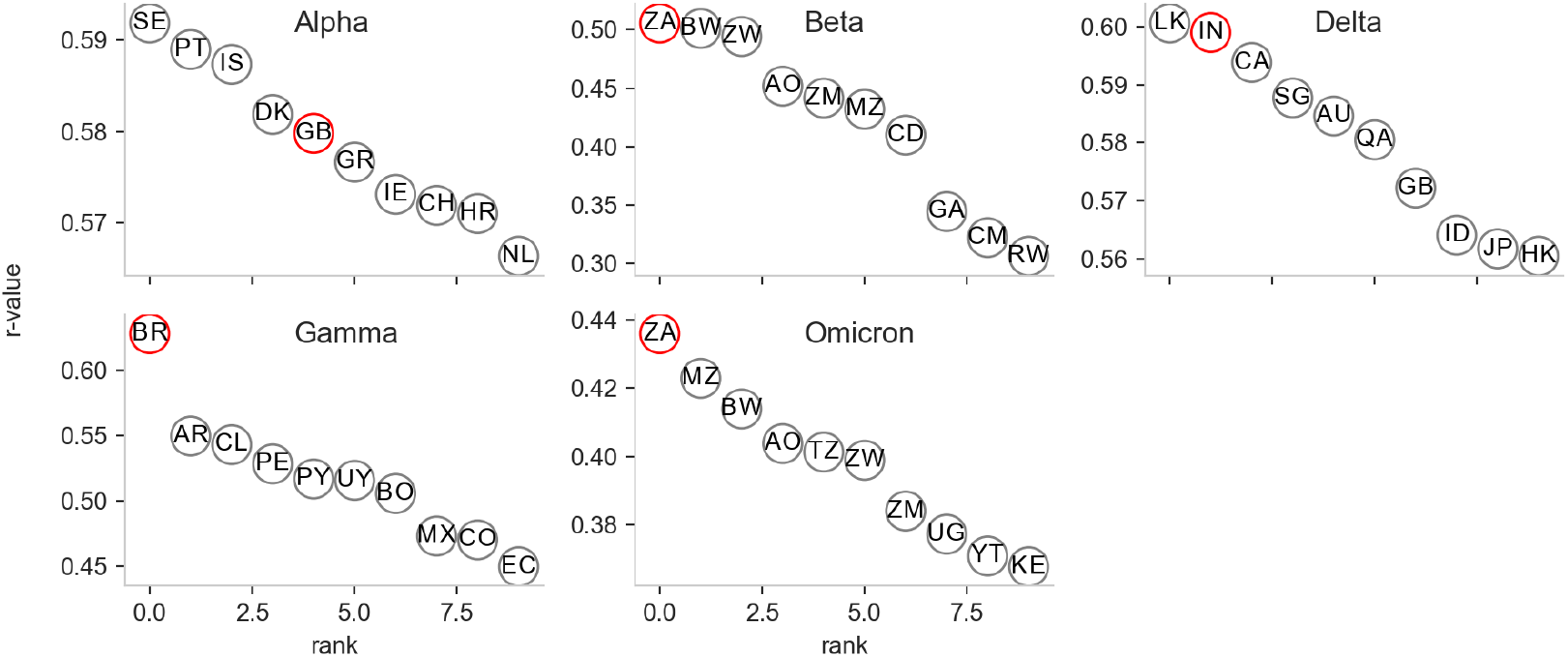
Arrival prediction (r-value) for the 10 best outbreak candidate. The r-value between the import risk distance *D*^*IR*^(*m*|*n*_0_) = − log(*p*_∞_(*m n*_0_)) and the arrival time for the 10 best ranked outbreak countries (*n*_0_). The 2 Letters in the circles are the countries ISO alpha-2 codes. The red circle marks the country declared as outbreak country by the WHO.

**Figure S7:**
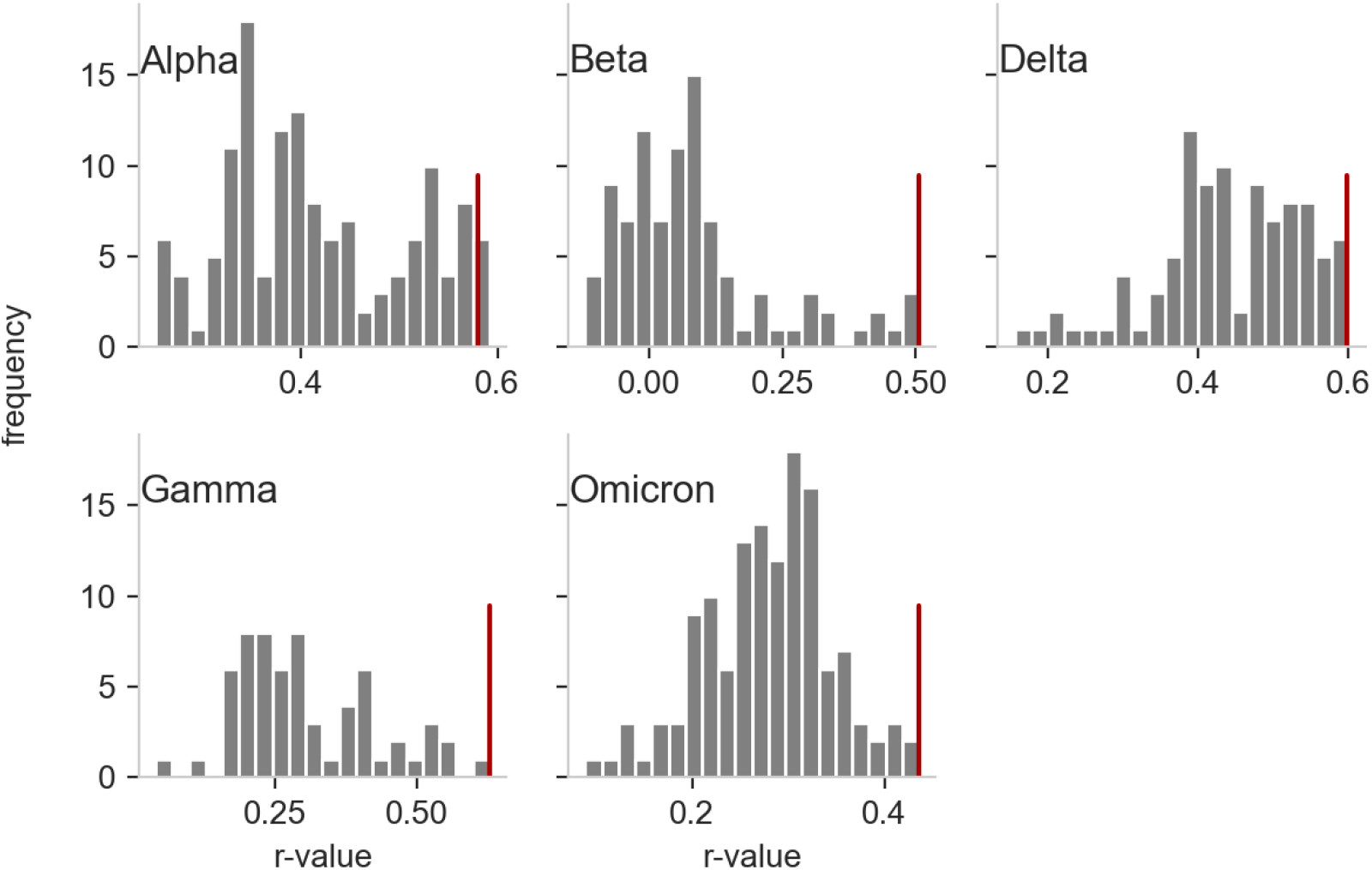
Arrival prediction performance (r-value) for the outbreak country candidates. The frequency of the r-value between the import risk distance *D*^*IR*^(*m*| *n*_0_) = − log(*p*_∞_(*m*|*n*_0_)) and the arrival time for all possible outbreak countries. The red vertical line marks the r-value using the country declared as outbreak country by the WHO.

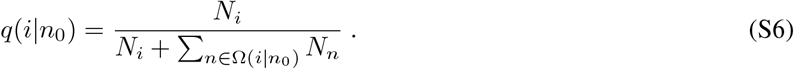

Now, we use the SPEx on a random walk that starts at *n*_0_ and the walker exits at node *i* with probability *q*(*i*|*n*_0_) or continues its walk with probability 1 − *q*(*i*|*n*_0_). Thus, the probability to be at node *m* if the walker was before at node *m* − 1 is

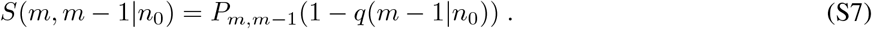

Consequently, the probability to take a path Γ starting at *n*_0_ and exiting at *m* is

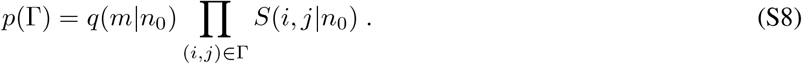

The probability to exit at node *m* from all possible paths (of all possible lenghts) is

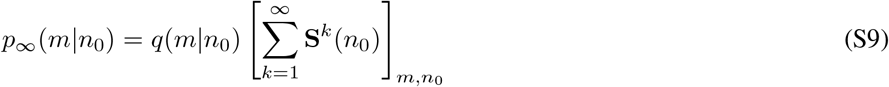

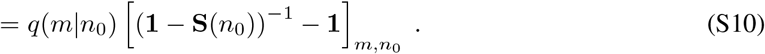

Note that 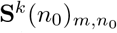 is the probability sum of all paths that started in *n*_0_ and end after *k* steps in *m*. We aggregate all airports of the same country by computing the weighted mean with weights

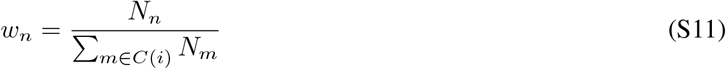

with *C*(*n*) as the set of airports that belong the same country as node *n* does.

##### II.3 Relation to distance and arrival time

In order to assess the quality of the import risk, we compare it with the arrival time of past variants. Clearly, the higher the import risk to a country, the earlier it is to arrive and the direct relation between the probability of travel to a city *m* from a city *n*_0_ and the mean first arrival time *t*_1_ is

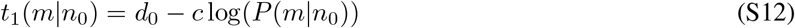

which is the effective distance [10, 11]. Thus, we define the import risk distance as

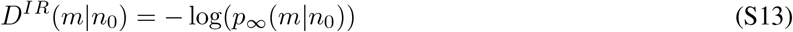

which is proportional to the mean first arrival time.

##### II.4 Alternative distance measures

There are alternative measures to estimate the arrival time [10, 12, 13], and we want to compare our import risk distance to these established measures. However, please note that the alternative measures have a clear qualitative relation to the arrival time, but it is not possible to directly infer the number of passengers that travel between airports from them (what the import risk is especially designed for). The already introduced alternative measure is the effective distance [10] that uses the flow between airports to estimate the probability to travel from airport *n* to *m*

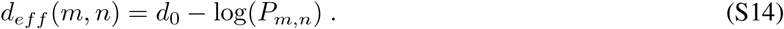

Now, the distance along a specific path Γ that connects *m* and *n*_0_ is the sum of the path elements distances

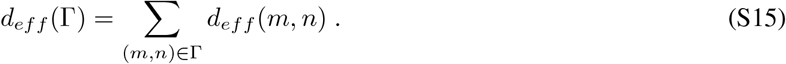

Finally the effective distance from airport *n*_0_ to *m*, also not directly connected airports, is the minimal effective distance of all possible paths Ω(*m, n*_0_) they are connected through

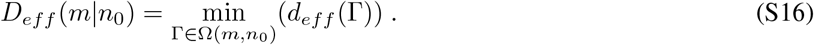

An extension to the effective distance is the random-walk effective distance [13] that considers all possible paths connecting two airports Ω(*m, n*_0_) instead of only taking the dominant path with the shortest distance:

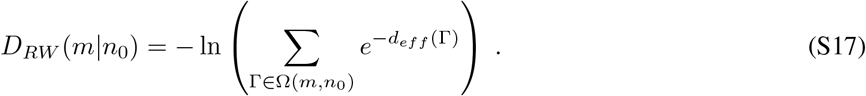

Note that the sum of path distances via their exponential is due to the linkage to the arrival time as explained in [13].

We also add a comparison with a metric derived from Diffusion Distance [12] which exploits the definition of a random walk Laplacian on top of the WAN. We further explain this Information Distance *D*^*ID*^ in the dedicated section V.

###### Country-Level aggregation

The country-level aggregation of the import risk distance *D*^*IR*^ is done by first aggregating the import risk on country-level (as described in Sect. II.2) and then applying Eq. S13.

To aggregate the other distances (*D*_*eff*_, *D*_*RW*_) we could either take (along the line of *D*_*eff*_) the minimal distance between two countries (of all relevant airport pairs), or use a weighted multipath approach as used in the derivation of *D*_*RW*_. We will highlight the latter in the following; however, we also computed the minimal measure and found that it is outperformed by the multipath distance (not shown, but it is the basic finding in [13]).

As shown in [11], the effective distance of two paths combined is

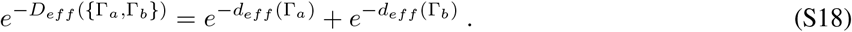

Thus, the multipath (MP) effective distance that considers all shortest paths from country *S* to *M* is:

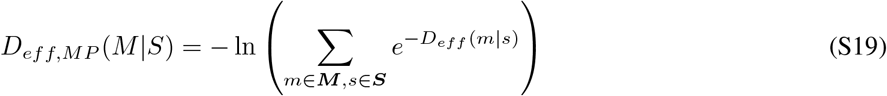

with ***M*** as the set of all target airports in country *M* and ***S*** all source airports of country *S*.

Since the distance of source airports with a larger population are more important, we additionally weight the source airport with *w*_*i*_ = *F*_*i*_*/* ∑_*s*∈***s***_ *F*_*s*_, which represents the probability of an infected to start in location *n*. Now, we compute the population weighted multipath effective distance by

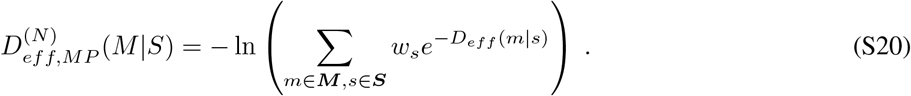

Note that the weighting for the effective distance can be reformulated to

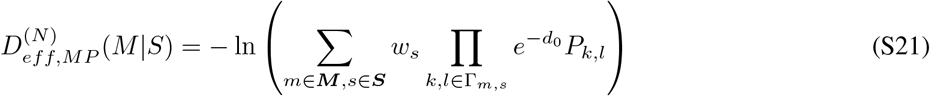

which corresponds to multiplying the probability to start at the source airport *s* to the first step of each path. Analogously the

##### II.5 Data for arrival time and outbreak region

We compare the import risk to measured arrival times of different variants. Therefore, we need to define the outbreak-country and -month and the arrival times. We defined these variables in different ways.

###### (I) external sources

Here we rely on peer reviewed [8] or official [14] sources. The outbreak country and the outbreak month are taken from the website of the World Health Organization (WHO) “Tracking SARS-CoV-2 variants”[14] and the arrival times of the variants Alpha, Beta, Delta, Gamma and Omicron were externally computed with “grinch”[8] and taken from their project website[7]. If arrival times are before the official outbreak they are removed from the analysis (for Delta=1, Gamma=1 and Omicron=19 countries are removed).

###### (II) GISAID data

To also use the other variants to validate our import risk method we design a simple arrival time algorithm. First, we need to define the outbreak day. Instead of relying on an official definition from the WHO, we use GISAID data. The outbreak time *T*_*X,out*_ of variant *X* is defined by

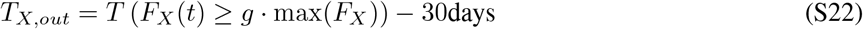

with *F*_*X*_ (*t*) being the fraction of variant *X* to all sequenced probes at time *t* and *T* (*F*_*X*_ (*t*) ≥ *g* · max(*F*_*X*_)) the time when *F*_*X*_ (*t*) crosses the first time the threshold *gcdot* max(*F*_*X*_) where *g* ∈]0, 1 [and we set *g* = 0.025. In other words, the outbreak is defined by 30 days before the variant reached 2.5% of its worldwide peak. We estimate the arrival time of variant *X* in an country by the most simple way: the first time the variant is detected (according to GISAID data). In Fig. S8 the estimated outbreak time, official WHO and arrival times of each country are shown. Since for some variants (Alpha, Delta, BA.2) many arrival times fall clearly before our estimated and even the official outbreak date, we recomputed for these countries the arrival time to the first GISAID-detection after the outbreak date. We argue that either (i) the sequencing of the variant in these countries was error-prone (1. count is very sensitive to any wrong detection) or (ii) the spreading was slow and the variant did not dominate the local epidemic until it reached a susceptible country (low NPIs) from where it did spread more easily (probably the case for Delta).

**Figure S8:**
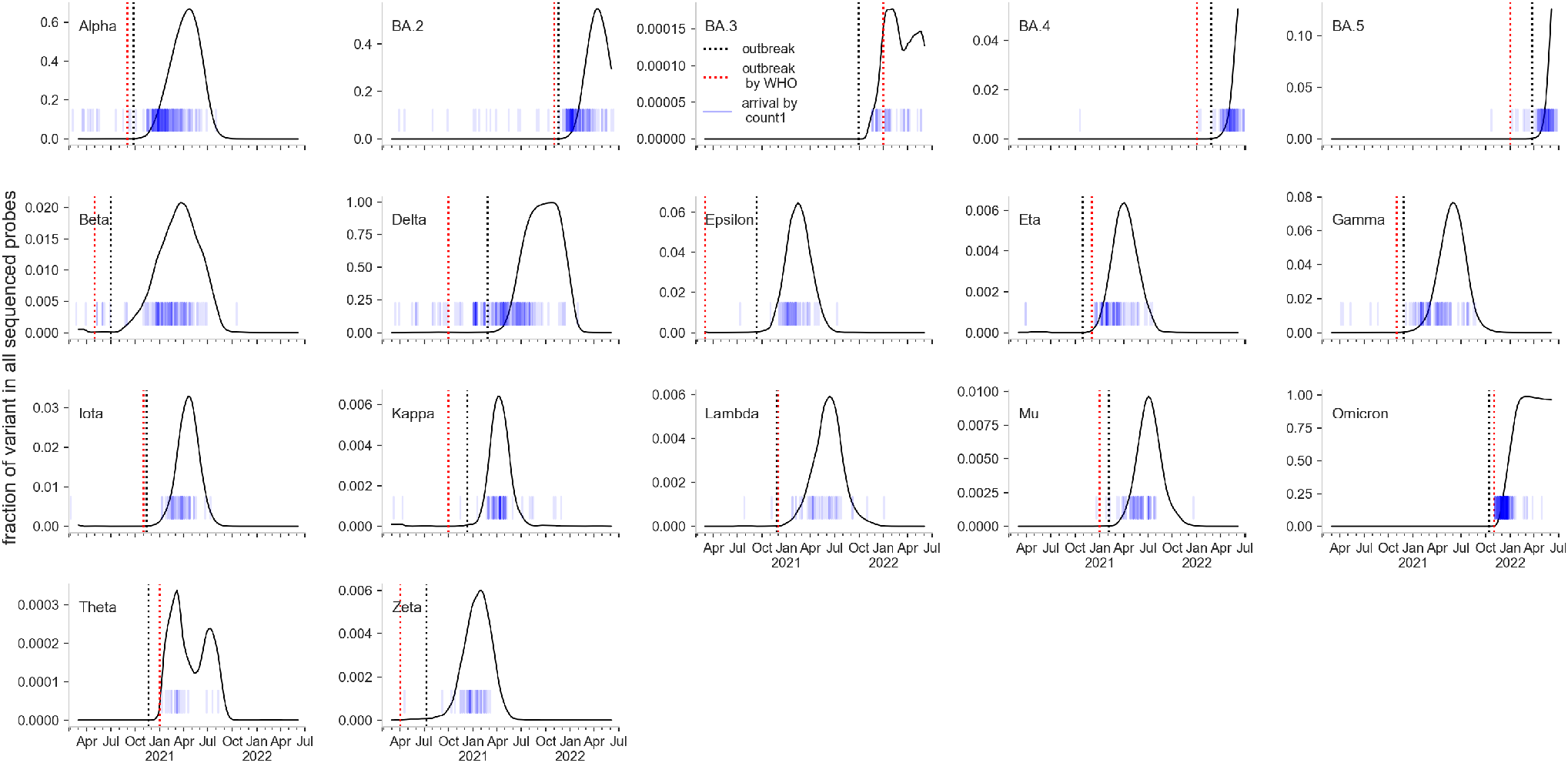
Outbreak defined by fraction of all sequenced probes. The outbreak date (black dashed vertical line) of a variant can be defined by the first time the fraction of a variant *X* of all sequenced probes reaches 2.5% of its current worldwide peak. To exclude maldetections of 1st. arrival times in countries, we exclude all arrival times (blue short vertical lines) that are before the outbreak date and set the arrival time as the first detection in the respective country after the outbreak date. The official outbreak date by WHO is marked by a red dashed vertical line.

##### II.6 Outbreak detection based on 1st count GISAID data

If we repeat the outbreak detection method using all variants and the arrival times estimated via GISAID data (arrival by first detection, Fig. S8), we see that the outbreak detection via the best correlation between import risk distance *D*^*IR*^ and arrival times *T*_*arrival*_ in general confirms the outbreak regions declared by the WHO (see Figs. S10, S9). There is a discrepancy for Delta. While using WHO and “cov-lineages.org” data, the official outbreak country India (IN) was second best, it is only on rank 12 if our GISAID estimates are used. A possible explanation is, that our outbreak date estimation is 5 months after the WHO date. In order to not lose the countries with arrivals before the outbreak date, we recompute the arrivals by the first count after the estimated outbreak date. One can argue that Delta did locally spread much stronger in South Africa (ZA, the top ranked country), and therefore is ZA for the worldwide distribution of larger importance than India. An alternative explanation is that the passenger flow in the WAN was too low and when it increased, ZA had a more active Delta epidemic.

**Figure S9:**
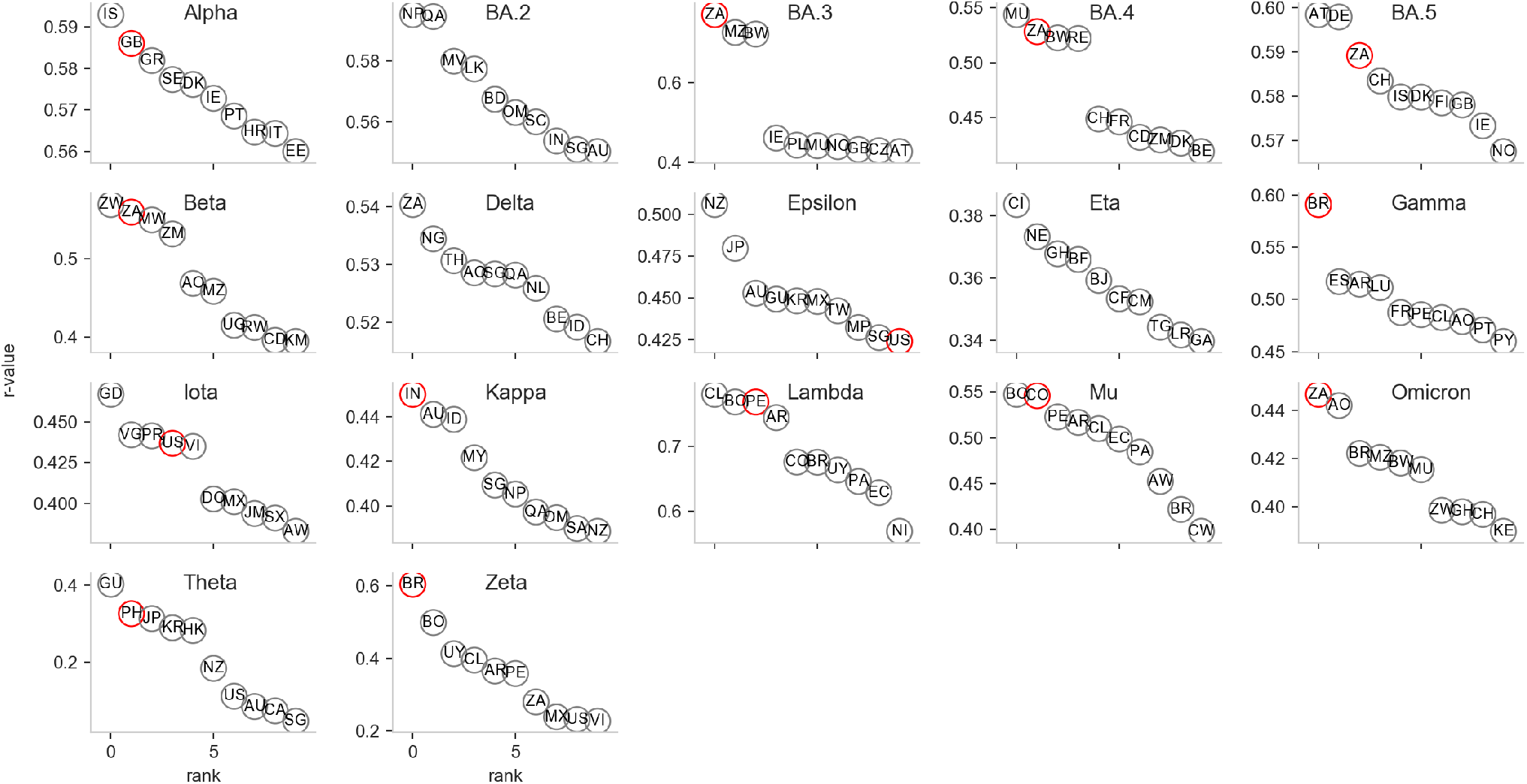
Arrival prediction (r-value) for the 10 best outbreak candidate. The r-value between the import risk distance *d*_∞_(*m*|*n*_0_) = − log(*p* (*m*|*n*_0_)) and the arrival time for the 10 best ranked outbreak countries (*n*_0_). The 2 Letters in the circles are the countries ISO alpha-2 codes. The red circle marks the country estimated as outbreak country based on GISAID arrival times. In contrast to Fig. S6: the arrival times and outbreak dates are estimated via GISAID data (arrival by first count, outbreak date by reaching the first time 2.5% of worldwide peak of the respective variant).

**Figure S10:**
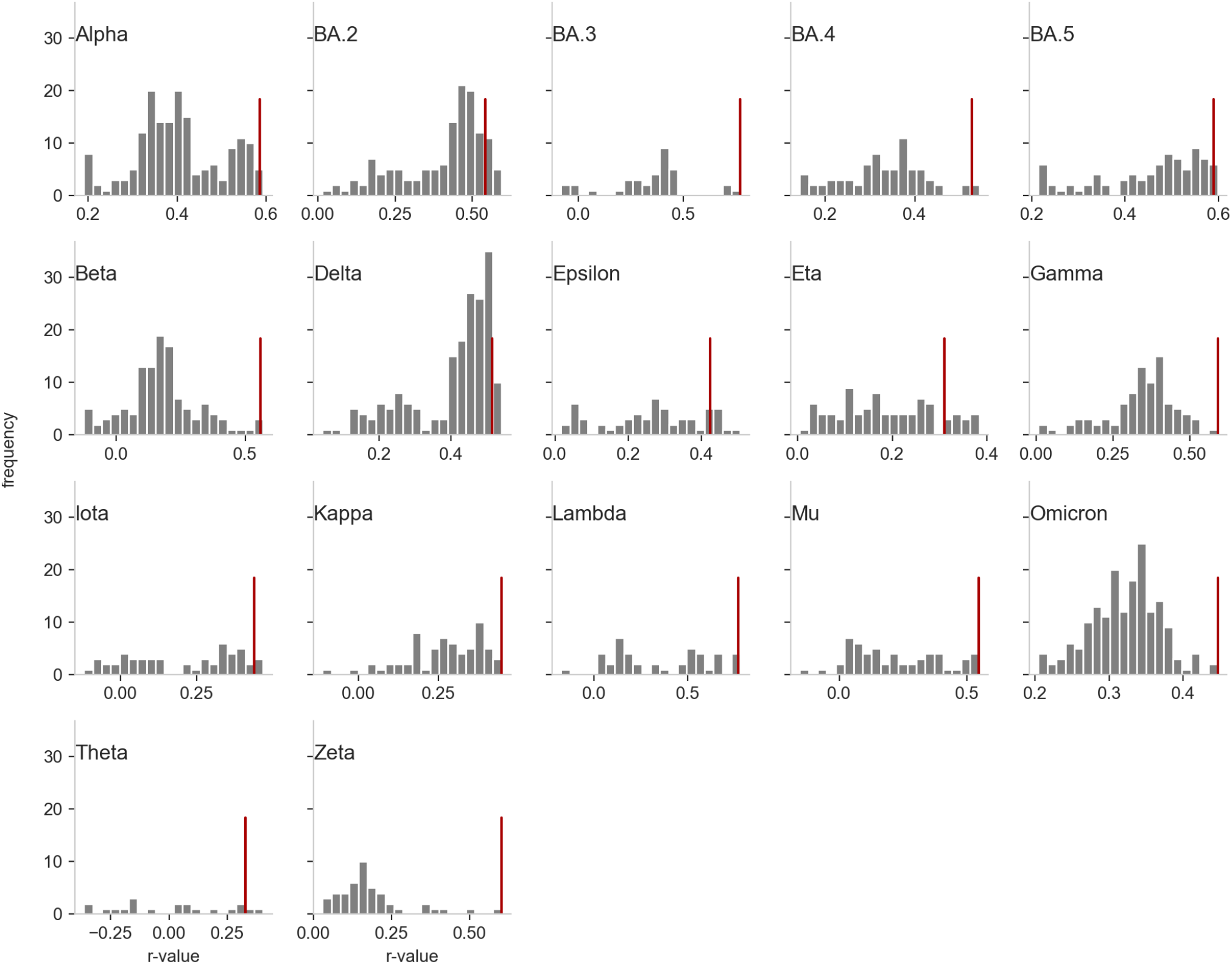
Arrival prediction performance (r-value) for the outbreak country candidates. The frequency of the r-value between the import risk distance *D*^*IR*^(*m*|*n*_0_) = − log(*p*_∞_(*m*|*n*_0_)) and the arrival time for all possible outbreak countries. The red vertical line marks the r-value using the country estimated as outbreak country based on GISAID arrival times. In contrast to Fig. S7: the arrival times and outbreak dates are estimated via GISAID data (arrival by first count, outbreak date by reaching the first time 2.5% of worldwide peak of the respective variant).

#### III. Epidemic Scenarios

We consider two distinct models to project the number of daily new infected people, namely, a renewal equation based model and a multi-strain SIR-like model. The first one is actually part of the pipeline, while the second one is used as validation.

##### III.1 Renewal equation

The renewal equation approach is a well-known technique, widely used in epidemiology [15, 16, 17]. The reason why renewal equations are such strong candidates for early projection of new cases, is the fact that informing them requires only the reproduction number of the new variant of concern, its generation interval distribution, and the number of people infected by the new variant who travel into the target country from the source country. This allows easily to explore scenarios with different values of epidemiological quantities of interest, such as the effective reproduction number of a new variant as it spreads from the source country to others through travelers.

For now, we assume that the susceptible population is much larger than the number of active cases, and that the mixing between the infected and the susceptible is homogeneous. This allows to exclude feedback loops in the dynamics, e.g. the fact that immunity to the new variant builds up through infection, which would modify the dynamics itself. Such strong assumptions are acceptable as long as we restrict our projections to the very first few weeks from the introduction of the new variant in the target country.

The model assumes that the number of newly infected people at day *t, I*(*t*), is given by two distinct processes: a) the arrival of infected individuals from the source country (*I*_*out*_(*t*)) and b) the daily new infections (*I*_*in*_(*t*)) happening in the target country due to the endogenous spreading. The former is estimated from section II, while the latter can be estimated through the renewal equation

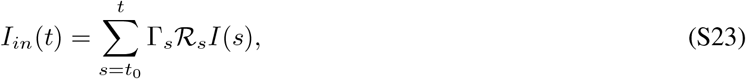

where *t*_0_ is the day the first infected cases arrived in the target country, ℛ_*s*_ is the daily reproductive number on day *s*, and Γ_*s*_ is the generation time distribution, i.e. the fraction of transmissions that would occur on day *s* after infection. Finally *I*(*t*) = *I*_*out*_(*t*) + *I*_*in*_(*t*). This is the simplest renewal process, which does not include the fact that the target population might have an inhomogeneous immunological landscape, due to previous infections or vaccination. To model this phenomenon, we reinterpret the term on the right side of equation (S23) as the number of inoculations spreading from currently infecting people, which will turn into infections depending on the susceptibility of the recipients. If we assume that previous infections (with other variants) protect against reinfection with an efficacy of *n*_*e*_, and, analogously, vaccination has an effectiveness of *ν*_*e*_, then we can explicitly account for removals by modifying equation (S23) into

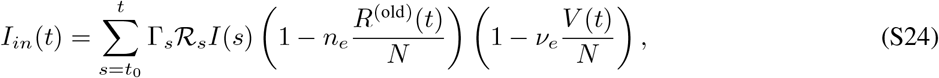

where *R*^old^(*t*) is the number of recovered people from previous variants that still have some protection against infections, and *V* (*t*) is the total number of vaccinated people. This assumes that the number of recovered or vaccinated people is uniformly distributed across the population, and that the events ‘being vaccinated’ and ‘having been infected’ are independent. This also assumes no gradual waning of protection against infection. However, we can consider as recovered or vaccinated only people who were infected or vaccinated recently, rather than from the beginning of the pandemic. For instance, considering only people who got either infected or their second dose up to six months prior to *t* is equivalent to assuming that there is an abrupt waning of efficacy against protection six months after getting infected or vaccinated.

Although these hypotheses might seem unrealistic, the lack of readily available data about waning and immunological landscapes of various countries, and the fact that this should be used only for short-term scenario explorations, allow us to avoid introducing further complexity into the model.

The cumulative number of cases and amount of fully vaccinated individuals at each day are the ones reported in the public repository at ^1^. We select the values for vaccine efficacy and protection from previous infection from available works. In particular we set the vaccine efficacy *ν*_*e*_ to 0 for Alpha, 0.5 for Delta, BA1 and BA2 and to 0.12 for BA.5 ([18, 19, 20, 21]). The selected protection against reinfection *n*_*e*_ is 1 for Delta, 0.56 for BA.1 and BA.2 Omicron lineages and 0.13 for BA.5 ([22, 23, 21]).

The second model is a multi-strain SIR inspired by [24]. This is a two-strain model in which people who recover after being infected with the former variant are not completely immune to infection from the latter variant. The equations governing this system are

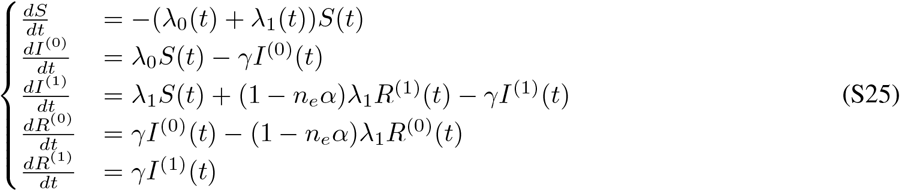

where 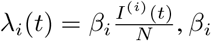 being the transmission rate of the variant *i*, and *γ* being the recovery rate. The initial condition 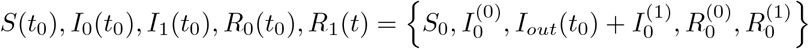. Note that, since *I*^*out*^(*t*) represents the arrivals from the source country at the beginning of each day, the system is not closed. This is not a problem because we are considering countries, so 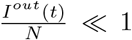. Since the dynamics does not include, per se, the fact that the initial condition changes every day due to arrivals, we can solve this system on a daily basis, updating the initial condition and restarting the system accordingly. The advantage of this system is that it includes feedback phenomena, which is good when validating the model, as it may need to run for more than a few weeks. The drawbacks are that informing the model requires good point estimates of the various compartments, and the interpretation of the transmissibility coefficient related to the measured ℛ_*t*_, which may not be straight-forward. For such reasons, this model is used to validate the renewal equation approach, in particular for countries where no new cases were observed after a few weeks from their emergence (not shown). Projections errors valuated with the SIR model relative to Alpha lineage are shown in

##### III.2 A fully worked out example: the Alpha variant

We apply our pipeline to a real case, the Alpha variant of concern (VOC), that was identified in the UK on 20 September 2020 [8]. We assume that the UK is the source country and we demonstrate how the pipeline works. In the following, we consider as the generation time interval distribution the one inferred from the literature [25].

Starting from the phylogenetic part of our pipeline, we take the time of emergence estimated when *n* = 20 sequences were collected, to simulate a realistic scenario where only little information is available. This gives a central estimate for the time of emergence of the Alpha variant around the 9^*th*^ of November 2020. The daily growth rate estimated is *r* = 0.097 (95% HPD: 0.008–0.202). To translate this into *R*_*t*_ in the source country, we assume that all the growth rate advantage of Alpha relative to the previous circulating variants is given only by transmission advantage (limited capacity of reinfections with Alpha). Further, typical generation time distributions are Gammas, as in [25]. This allows us to estimate the *R*_*t*_ using formula 2.2 in [26]:

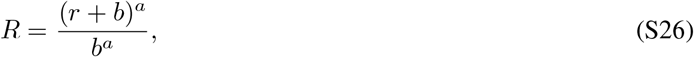

where *b* and *a* are the shape and rate of the Gamma distribution generation time. In our case, *a* = 5.9, *b* = 1.13, therefore *R*_*t*(*α*)_ = 1.62(1.04, 2.63).

For any target country, the projection of the number of cases infected with Alpha in the next weeks is performed in two steps: first, we estimate the number of infected travelers (referred to as seeds) who arrive in the target country from the source country, then we use the renewal equation (S24) on each possible scenario, to account for endogenous transmission of the secondary cases in the target country. The first step consists in using the import risk estimates described in section II to compute the number of daily travelers from source country to other target countries. We use import risk probability from source to target times the average daily outflow of passengers from source country using WAN data. We then determined the number of travelers infected with Alpha. This is done by considering the proportion of sequenced cases that are Alpha times the 7 − *day* moving average of daily incidence of new cases, assuming that sequences are taken randomly from the infected population. This estimate does not include undercounting in the source country, which we can estimate as follows.

For a given country, we use the daily new estimated COVID-19 infections from the IHME model, which is a hybrid with two main components: a statistical “death model” component produces death estimates that are used to fit an SEIR model component ^2^. For a complete overview of this model and a comparison with other estimates, we refer to OWID^3^. The data we used for our estimation are publicly available^4^. In a given temporal window, we integrate over time the confirmed number of cases (7d moving average) and the estimated true number of cases, as well as the estimates for its lower and upper bounds defining the 95% uncertainty interval. The mean undercounting factor is estimated by the ratio between the integrated estimate of the true cases and the confirmed ones in the temporal window, and similarly we estimate the corresponding uncertainty interval. We show in Fig. SS11 the undercounting factor obtained for all countries for which the data is available, whereas Fig. SS12 shows the evolution of this factor along periods of 6 months for some representative countries.

**Figure S11:**
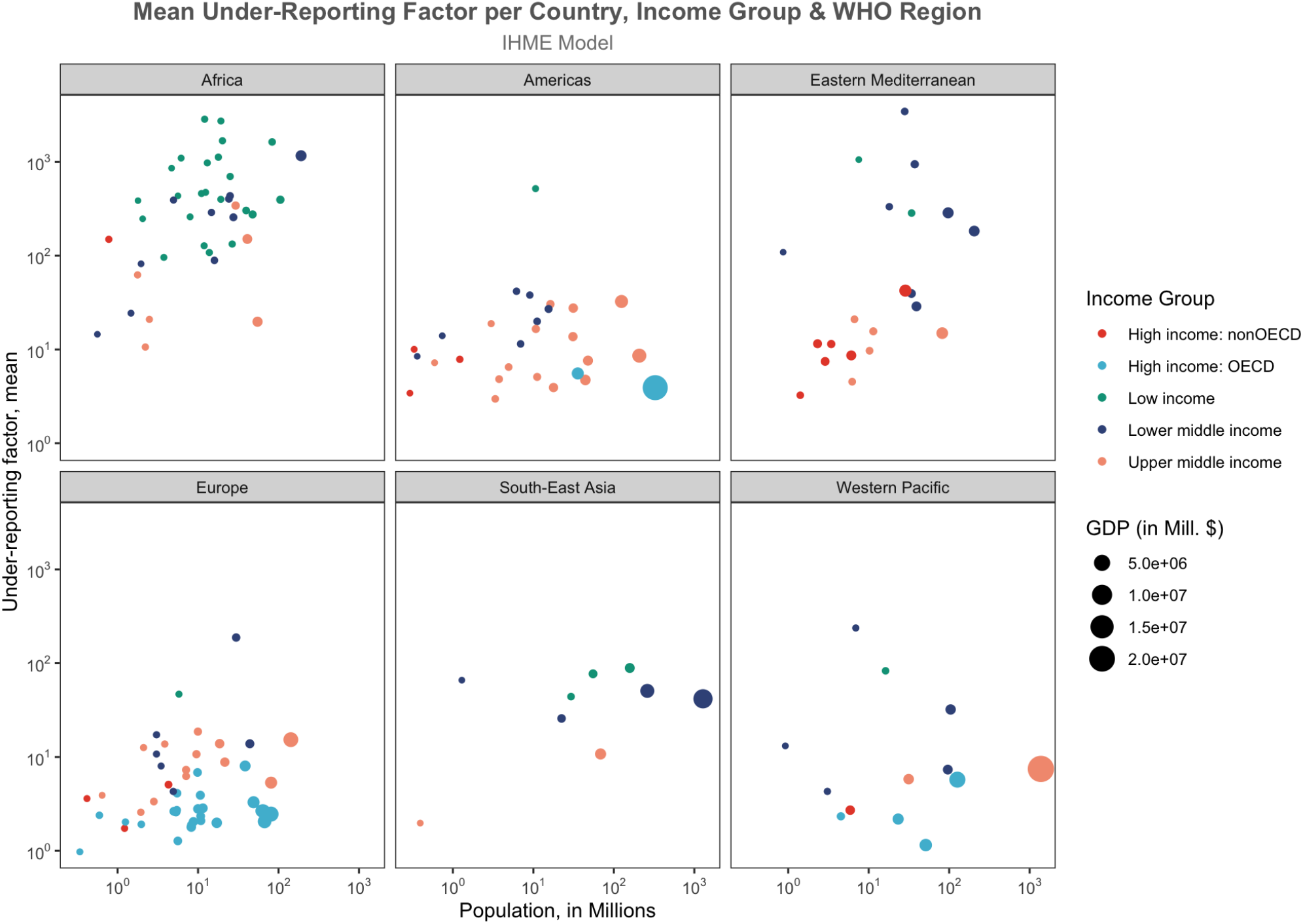
Undercounting factors by WHO region and income group. Estimates of the factor accounting for missing confirmed cases: values larger than 1 indicate that a country is counting and confirming less COVID-19 cases than the real number. The reference period is the first semester of 2022. See the text for further details.

To allow for variability in undercounting, we consider two extreme scenarios: the best one, where undercounting is assumed to be 2.27, and the worst one, where undercounting is assumed to be 2.97. The number of infected travelers from the source country to the target country is then computed by multiplying the number of travelers into the target country by the proportion of infected people in the source country. This is often not a natural number. This is not a problem, as the renewal equation does not need to use integer number of infected people, and we interpret this as the results of the various averaging performed through all the steps. The model produces the total number of infected people in the target country given the seeds and the ℛ_*t*_ by day of infection. To validate the model, we need to estimate how many people infected with the VOC were present in the target country during the considered period. We do so in the same way we estimate prevalence in the source country: by multiplying the proportion of sequenced cases that turned out to be Alpha times the daily incidence in the target country, scaled by the estimated undercounting factor.

**Figure S12:**
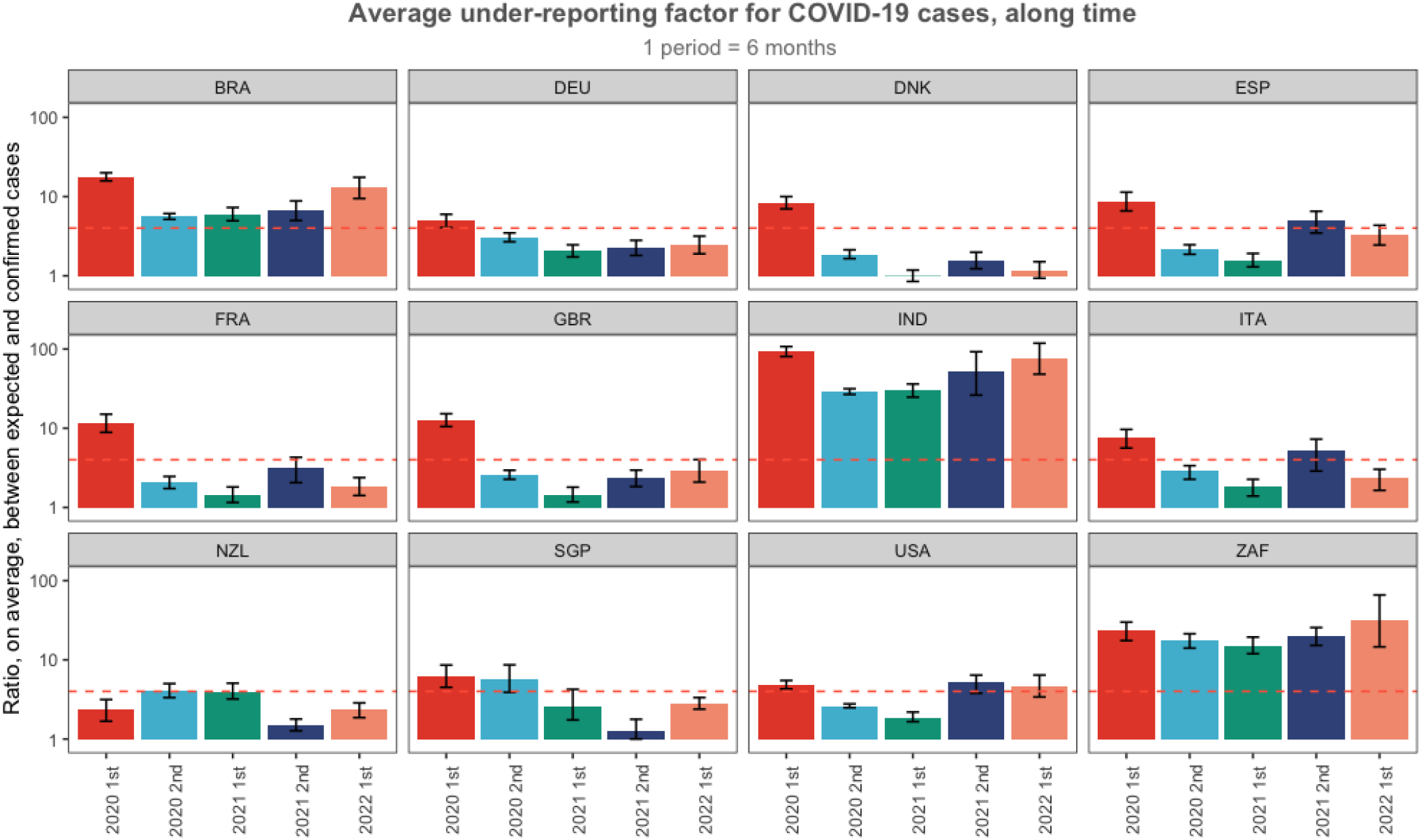
Undercounting factors over time. Estimates of the factor accounting for missing confirmed cases as in Fig. S11, where each panel describes the evolution along periods of 6 months for some representative countries. The dashed line indicates the value 4. See the text for further details.

The total number of different scenarios computed is, in this case 2 × 2 ×3: undercounting in both the source and the target countries, and the different reproduction number of the VOC. Results are shown in Figure 3C and in Figure S13A.

##### III.3 Prediction error

For each lineage we evaluate different scenarios with a) low and high values of underreporting in both source and target country b) three different basic reproduction numbers *R*_*t*_ that correspond to the range of growth rate values estimated from the phylogenetic reconstruction.

We infer from data the number of infected individuals with the emerging lineage in the target country *m* and we evaluate the prediction error as zero if this estimated number is included in the range identified by different epidemic scenarios. If the number of infected people evaluated from data is out of the range spanned by the epidemic curves, then the prediction error is evaluated as the root-mean-square error, normalized to the range of the data observed in the target country *m*, between observed and the closest simulated epidemic curve:

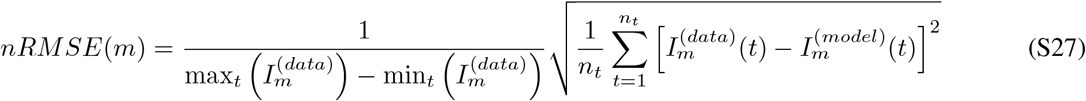

where *n*_*t*_ is the number of weeks with number of sequences greater than zero for the selected lineage in the considered country *m*, that is *n*_*t*_ is the number of available data points with not null infected people. Since the scenario simulations stop at the 3 week after sequencing was reported in country *m, n*_*t*_ is always *n*_*t*_ = 2. The idea behind the normalization by the data range is that it reflects the noise of reported sequences, i.e. if the sequencing rate is low, we expect a large variation and the sequencing data is less reliable. Prediction errors evaluated for all the considered lineages are shown in Figure 3 of the main document. All the panels report the nRMSE in each country as a function of both the number of daily passengers normalized to the total country population (x-axis, values for 100000 individuals) and the number of total collected daily sequences normalized to the total number of confirmed cases (y-axis, values for 100000 cases).

**Figure S13:**
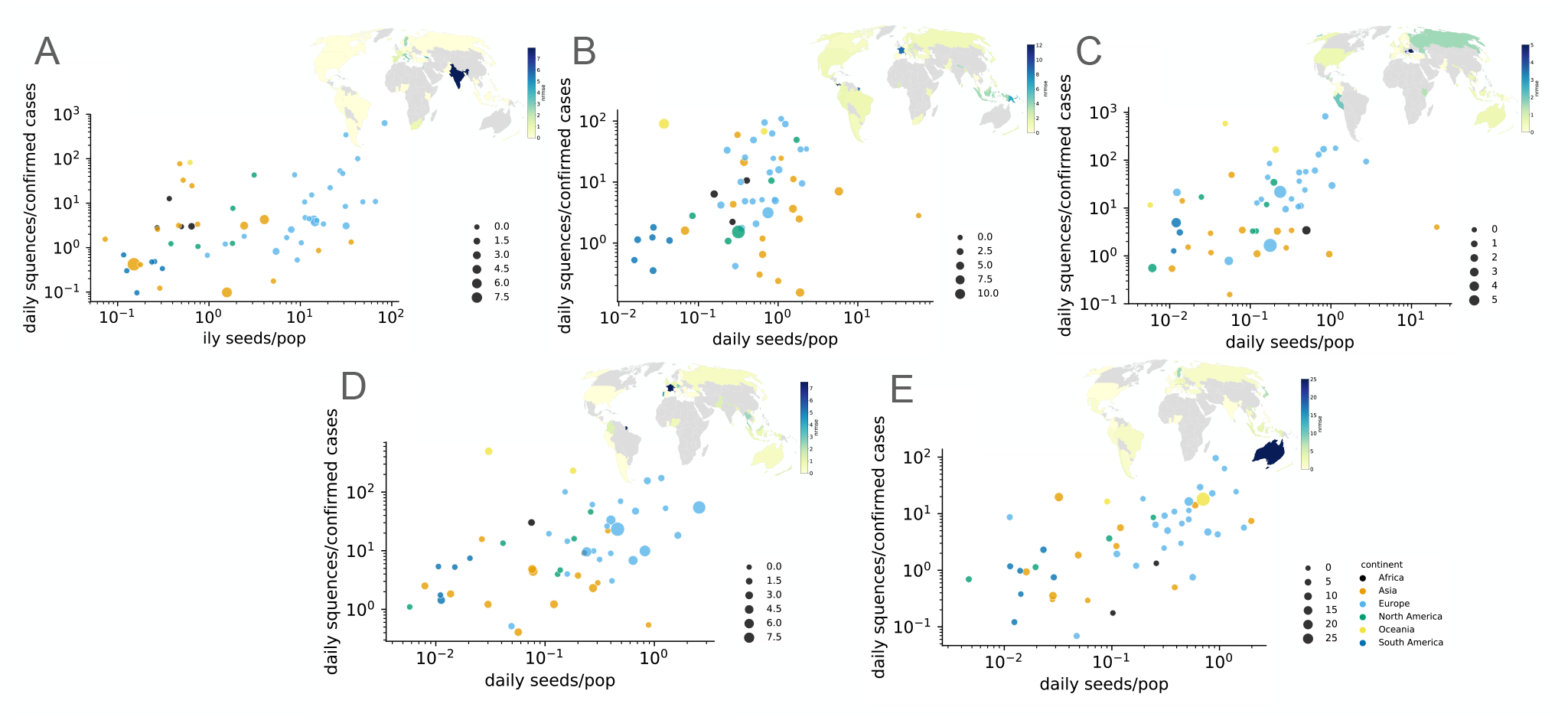
Epidemic prediction errors. Estimated errors between the number of individuals infected with an emerging lineage and the epidemic curves simulated in the considered scenarios. X-axis show the number of daily passengers normalized to the population in each country (for 100, 000 individuals), y-axis report the number of collected daily sequences, without any classification per lineage, normalized to the total number of confirmed cases (for 100, 000 cases). Inset panels show the map of prediction errors in each country. Panels A-E refer to, respectively, Alpha, Delta, BA.1, BA.2 and BA.5 lineages.

Insets show the evaluated error in each country. Results assess that, in most of the country, the simulated scenarios encompass the data and the prediction error is evaluated as zero. Moreover, error values greater than zero can be found for countries with higher passenger flows.

**Figure S14:**
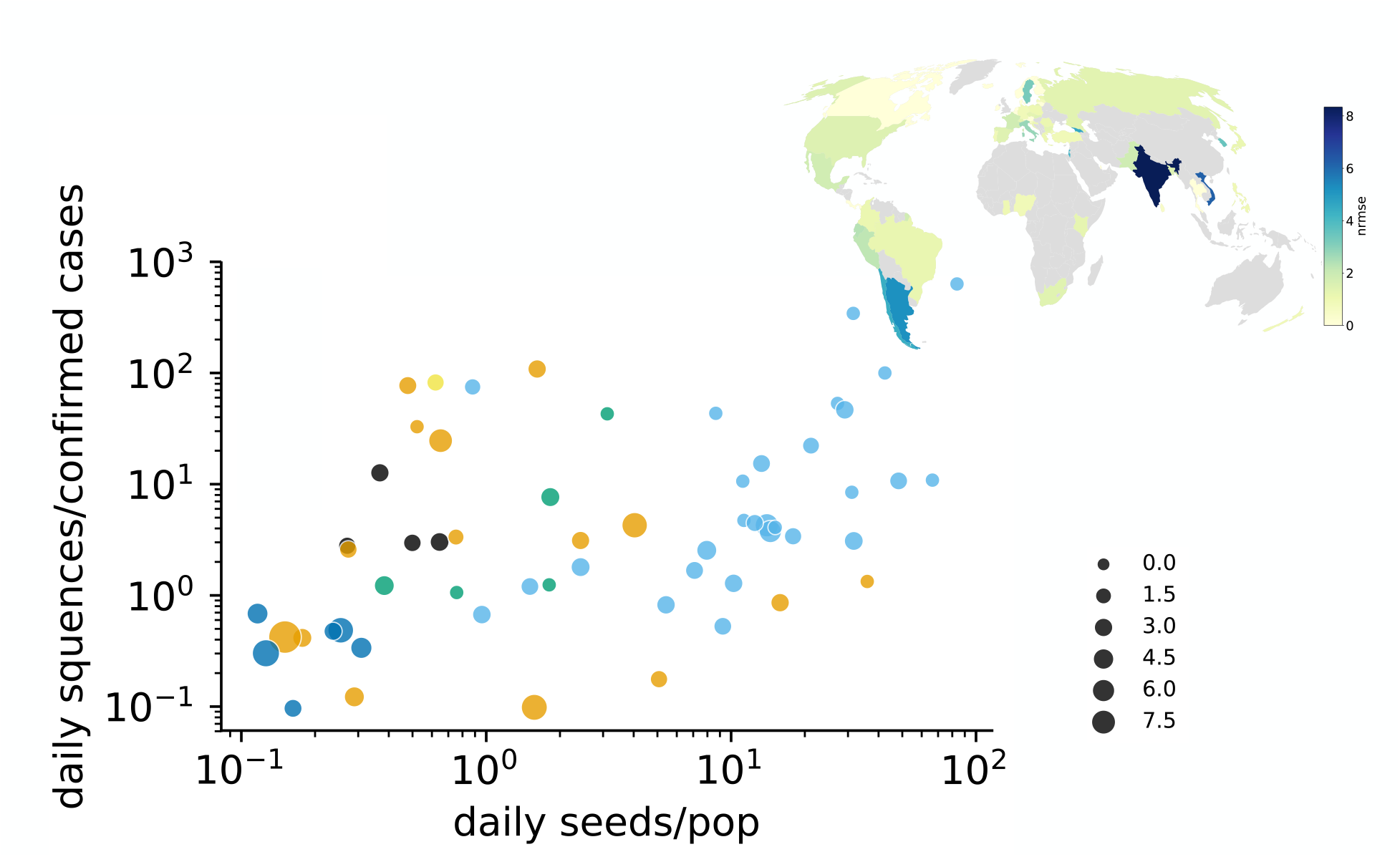
Epidemic prediction errors with SIR model, Alpha lineage. Estimated errors between the number of individuals infected with an emerging lineage and the epidemic curves simulated in the considered scenarios. X-axis show the number of daily passengers normalized to the population in each country (for 100, 000 individuals), y-axis report the number of collected daily sequences, without any classification per lineage, normalized to the total number of confirmed cases (for 100, 000 cases). Inset panels show the map of prediction errors in each country.

#### IV Pandemic delay

The pandemic delay estimates the time needed since tMRCA for a specific variant to reach a certain percentage *y* in a target country. It depends in general on a large variety of factors as the reproduction number, the fraction of vaccinated, the variant’s immune escape, season, weather conditions, the number, duration and strength of active non-pharmaceutical interventions (NPI), the national and international mobility and the epidemic situation. In the following estimation of the pandemic delay, we assume that the main driver/predictors for the pandemic risk are the international mobility, the effective reproduction number and the country specific epidemic situation.

We will use a simple framework to combine the measures that is based on the replicator equation [27], stating that the fraction of a new variant can be described by a simple logistic growth equation (illustrated for Delta lineage in Figure S15). It assumes that there are 2 competing populations, the mixed population of all preexisting variants of size *N*_*pre*_ and the population of the emergent variant *N*_*x*_. According to the replicator equation, the evolution of the fraction *x* of the new variant in the whole population corresponds to

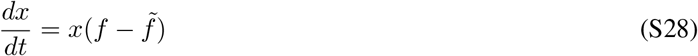

with *f* as the fitness of the new variant *x* and 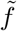 as the mean fitness, i.e.

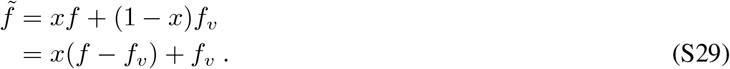

We can therefore rewrite the time-evolution to

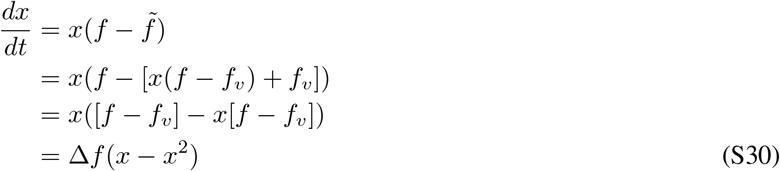

that has the logistic function as general solution

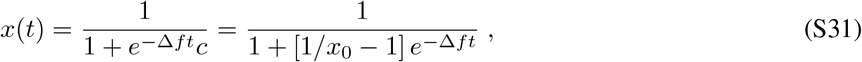

with *x*_0_ as initial condition being the imported infected cases from the country of origin *n*_0_ to the target country *m*

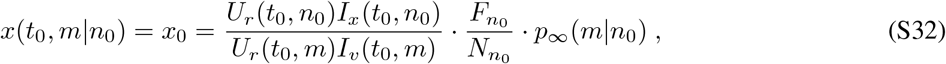

with *t*_0_ = tMRCA, *U*_*r*_ (*t*_0_, *m*) as the underreporting factor of cases in country *m* (introduced in Sec. III.2), 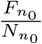 as the probability of leaving the country via the WAN and *p*_∞_(*m*|*n*_0_) as the import risk (see Sec. II). Note that with Eqs. S31, S32 we assume that the initial import *x*_0_ dominates, i.e. imports at later times can be neglected (otherwise a constant flux needs to be implemented). The fitness difference between the new variant vs. the already existing variant mix is approximated by

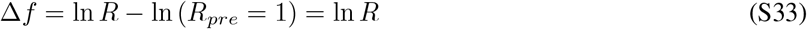

i.e. we assume that the reproduction number of the preexisting variant mix is one, motivated by the observed fluctuations around *R*_*pre*_ = 1 due to the behavioral and/or medical adaptation to the local epidemic situation.

The pandemic delay *t*_*y*_ is the time needed for the new variant to reach the fraction *y* of the infected population, where *t*_*y*_(*m*) for a specific country *m* is (rearranging Eq. S31)

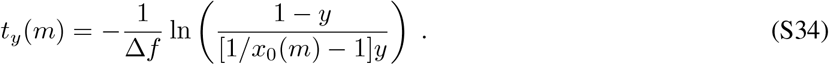

We can further simplify the pandemic delay by assuming that the initial import is small

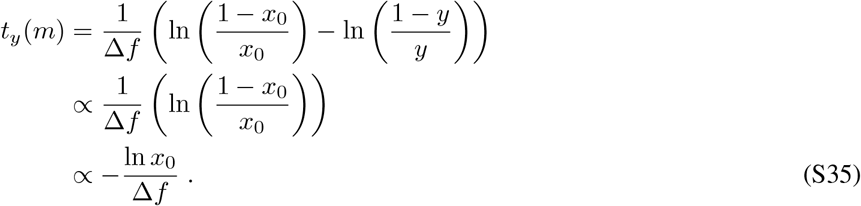

However, this simplification is merely meant as a help to ease understanding of the functional relations. In the manuscript, we use explicitly Eq. S34.

**Figure S15:**
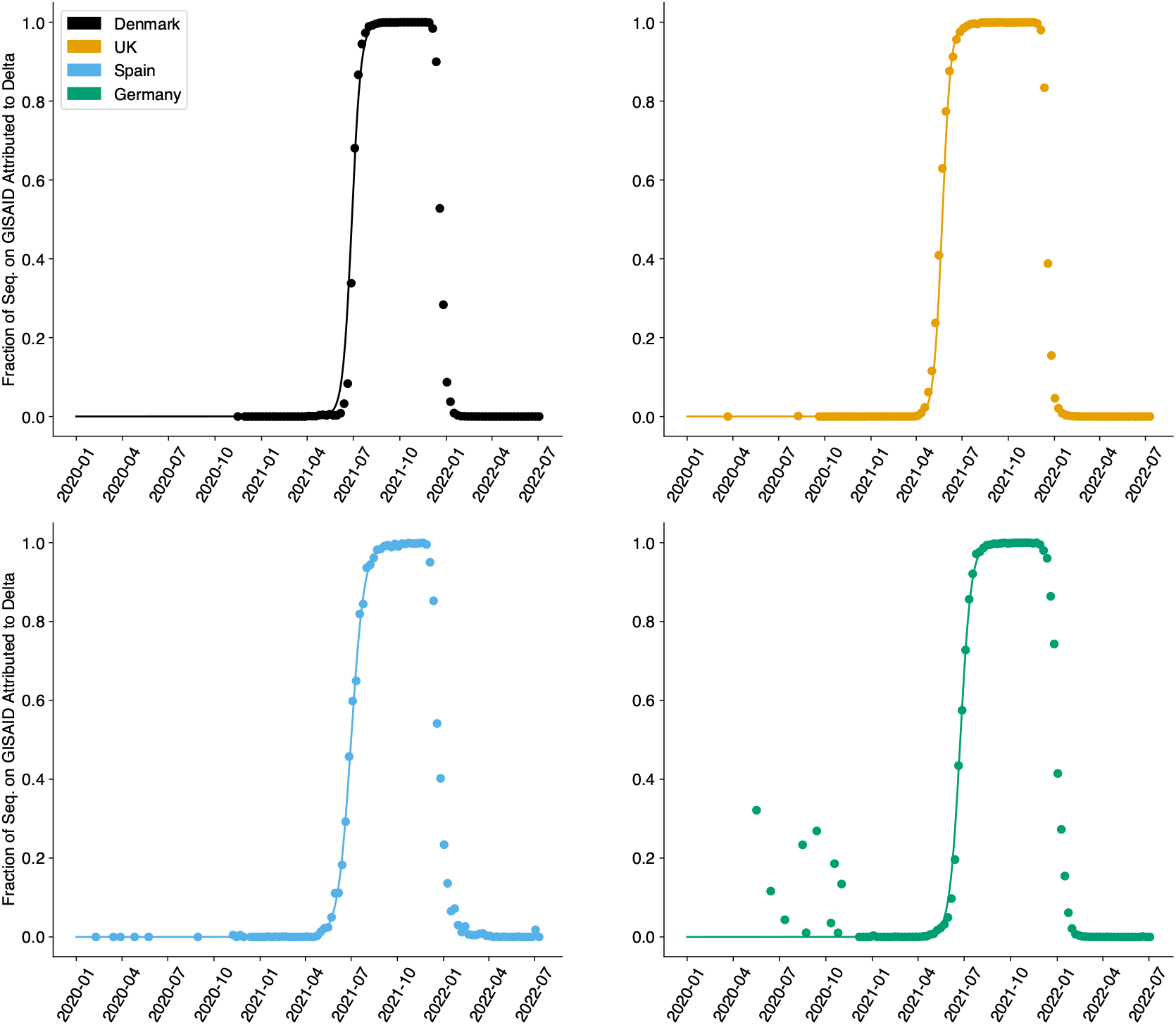
The fraction of seq. on GISAID attributed to the Delta variant for four example countries. As described for the Alpha variant by Fort [27], the relative fraction of a new variant can be accurately described by a simple logistic growth equation (Eq. S31).

#### V Information Distance

We also devise an alternative definition of distance on top of a network which embeds information from multiple-pathways diffusion as an additional comparison to the import risk measure. Distances based on the diffusive properties of the system have been of interest in recent years [10, 13]. Another key example is the Diffusion Distance [12] which estimates a metric distance between nodes based on how similarly the random walkers explore the network by using those nodes as sources, under the assumption that a mesoscale structure is recovered during the time scales in which the random walker explores its functional community.

Starting from Diffusion Distance definition, we propose an educated rewrite of the measure that fits the problem under study to predict arrival times of a random walker on the network, such as an infectious traveler from a source country. The probability **p**(*t* | *i*) of a walker to be in any point in the network at time *t*, starting from node *i*, embeds information of multiple paths via successive applications of the Laplacian operator. We introduce a new measure that merges this concept from Diffusion Distance and also embeds information from Effective Distance [10], namely, the idea that low probabilities *p*_*k*_(*t* | *i*) are associated with large distances. This can be embedded by taking the negative of the logarithm of the probability, in analogy with Shannon’s entropy. We now introduce this candidate measure for diffusive dynamics which we define Information Distance:

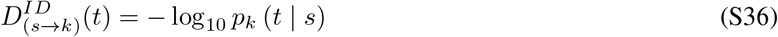

in which *p*_*k*_ (*t* | *s*) represents the *k* − *th* entry associated with node *k* of the probability state 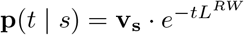. Here **v**_**s**_ is the initial condition probability for the walker starting from node *s*, the canonical vector with *s*-th component equal to 1. The random walk normalized Laplacian (*L*^*RW*^) [28] term encodes the probability to move from node *i* to node *j* in its matrix elements. Its off-diagonal terms can be computed as the negative value of *P*_*ij*_, which is directly estimated from the WAN weighted links as stated in subsection II.2. Given the multiple timescales involved in this definition, we evaluate the metric at different scales *t* to find the timescale at which *D*^*ID*^(*t*) performs better.

**Table S1:** Sequencing rates in the outbreak countries (Source) of SARS-CoV-2 B.1.1.7 (Alpha), B.1.617.2 (Delta), B.1.1.529 (BA.1), BA.2, BA.5 and BA.2.75 (Omicron) lineages. The outbreak countries (Source) are represented by their ISO alpha-3 codes (GBR: Great Britain, IND: India, ZAF: South Africa). The naive sequencing rate (Naive Seq. Rate) was computed by the ratio between new weekly cases (based on OWID-data [55]) and the weekly collected sequenced samples (based on GISAID-data [32]). We compute the final sequencing rate (Seq. Rate) by dividing through the underreporting factor (Underrep. Fact.) whose estimation is described in Sec. III.2. Both estimates are averaged for the lineage respective time-period between the median time of the most recent common ancestor (tMRCA) and the time when the first 50 samples got collected (*t*_50*S*_).

| Lineage | Source | tMRCA | $t_{50S}$ | Underrep. Fact. | Naive Seq. Rate [%] | Seq. Rate [%] |
| --- | --- | --- | --- | --- | --- | --- |
| Alpha | GBR | 13 Sep 2020 | 1 Nov 2020 | 2.6 | 0.19 | 0.075 |
| Delta | IND | 30 Aug 2020 | 7 Feb 2021 | 28.9 | 2.49 | 0.086 |
| BA.1 | ZAF | 31 Oct 2021 | 5 Dec 2021 | 19.7 | 51.8 | 2.623 |
| BA.2 | ZAF | 24 Oct 2021 | 19 Dec 2021 | 19.7 | 1.1 | 0.056 |
| BA.5 | ZAF | 16 Jan 2022 | 17 Apr 2022 | 31.6 | 2.94 | 0.093 |
| BA.2.75 | IND | 10 Apr 2022 | 12 Jun 2022 | 76.4 | 0.41 | 0.005 |

## Footnotes

1 https://ourworldindata.org/

2 https://covid19.healthdata.org/

3 https://ourworldindata.org/covid-models.

4 https://ourworldindata.org/grapher/daily-new-estimated-covid-19-infections-ihme-model.

